# Improving risk analysis of the environmental drivers of the spillover, emergence/reemergence, and spread of Crimean-Congo haemorrhagic fever virus, Marburg virus, and Middle East respiratory syndrome coronavirus in the East Africa Region

**DOI:** 10.1101/2024.11.07.24316934

**Authors:** Oluwayemisi Ajumobi, Meghan Davis, Christine Marie George, Lori Rosman, Sophie von DobSchuetz, Crystal Watson, Jennifer Nuzzo

## Abstract

**Introduction:** Emerging and/or reemerging infectious diseases (EIDs) in the East Africa region are associated with climate change induced environmental drivers. There is a need for a comprehensive understanding of these environmental drivers and to adopt an integrated risk analysis (IRA) framework for addressing a combination of the biological, environmental, and socio-economic factors that increases population vulnerabilities to EID risks, to inform biological risk mitigation and cross-sectoral decision-making. The aim of this integrative review was to identify knowledge gaps and contribute to a holistic understanding about the environmental drivers of Crimean-Congo haemorrhagic fever virus (CCHFV), Marburg virus (MARV), and Middle East respiratory syndrome coronavirus (MERS-CoV) infections in the East Africa Region, to improve IRA processes at the environment-animal-human exposure interface

**Methods:** An integrative review search was carried out to identify relevant studies and reports from 2000 to 2024. Searches were conducted in bibliographic databases and global institutional websites. Inclusion criteria were studies and reports (in English) addressing environmental drivers of CCHFV, MARV, and MERS-CoV infections across countries in the East Africa region, existing risk frameworks/methodological tools, and/or One Health policy recommendations for risk analysis of environmentally driven biological threats.

**Results:** Of the total number of studies retrieved from database searches (N = 18075) and website searches (N= 44), 242 studies and reports combined were included in the review with the majority covering the environmental drivers (N = 137), the risk frameworks/methodological tools (N=73), and the policy recommendations (N = 32). We identified ten categories of environmental drivers, four thematic groups of risk frameworks, and three categories of policy recommendations. Overall, many of the included records on the risk frameworks/methodological tools expounded on the adoption of ecological niche modeling (ENM) for environmental monitoring of potential transmission pathways of EIDs and other biological threats.

**Conclusion:** This integrative review recommends the adoption of specialized risk mapping approaches such as ENM for environmental monitoring of EIDs under IRA processes. Findings from the review were used for the conceptualization of an IRA framework for addressing environmentally driven EIDs.

**What is already known:** Outbreaks of emerging and/or reemerging infectious diseases (EIDs) have been increasing in frequency and severity over the last few decades and the risk of severe outbreaks are further aggravated by climate change and other anthropogenic factors.

**What this study adds:** Our review study adds to knowledge about context-specific environmental drivers and other related cascading risk factors that facilitate the transmission of EIDs with epidemic and pandemic potential in the East Africa region. In recognition of the need to address gaps in existing risk assessment processes, findings from the review were used for the conceptualization and initial development of an integrated risk analysis (IRA) framework that accounts for environmental drivers of pathogen spillover, disease emergence and spread across the entire spectrum of risk analysis; and to provide practical policy recommendations for enhancing the integration of the environment sector in the implementation of risk-based One Health interventions.

**How this study might affect research, practice or policy:** This study highlights the importance of incorporating environmental monitoring techniques under IRA processes to fill surveillance data gaps and to inform risk-based decision making for pandemic prevention, outbreak preparedness, and biological risk reduction.

## INTRODUCTION

Since the turn of the century, there has been a reported increase in the frequency and severity of infectious disease outbreaks and a heightened risk of occurrence of zoonotic emerging and/or reemerging infectious disease (EID) outbreaks.^1,2^ Environmental drivers like climate change (including variations in temperature and precipitation) and extreme weather events (EWEs) such as droughts and floods are reported to have an impact on the spread of infectious disease pathogens by increasing their survival patterns and geographic reach to new non-endemic areas.^3,4^ Countries in the East Africa region are experiencing increases in climate-induced EWEs and consequent infectious disease outbreaks including those of zoonotic origin.^5,6^ Of relevance to this study are three zoonotic EID pathogens included under the World Health Organization (WHO) priority Research and Development (R&D) blueprint list of pathogens with epidemic and pandemic potential:^7^ (i) Crimean-Congo haemorrhagic fever virus (CCHFV); (ii) Marburg virus (MARV); and (iii) Middle East respiratory syndrome coronavirus (MERS-CoV).

The severe public health threat posed by CCHFV, MARV, and MERS-CoV have the potential to result in global catastrophic biological risks (GCBRs) like the coronavirus disease 2019 (COVID-19) pandemic. GCBRs are described as largescale, high-impact biological risks with the potential to cause widespread disaster beyond the collective ability of national and international governments and private entities to control.^8^ This threat is further amplified by the impact of climate change and other environmental drivers, unavailability of vaccines or treatment options, health inequities among different population groups, and increasing evidence of the ability and/or potential to sustain human-to-human transmission.^9,10^ There are also increasing evidence of pathogen emergence/reemergence and seroprevalence reported across countries in the region [Supplemental Table 1 (ST1)].

### Risk Analysis

Lessons learned from past zoonotic pathogen spillover events and the resulting disease outbreaks, including outbreaks of severe acute respiratory syndrome (SARS) and Middle East respiratory syndrome (MERS), emphasize the need to enhance the detection of and response to EID outbreaks at early stages of pathogen emergence by having a better understanding of the ecology of EIDs and their origins in wildlife.^11^ This expanding knowledge calls for the adoption of integrated data-driven risk analysis approaches that properly account for the environmental drivers of EID events, to inform the establishment of early warning and collaborative surveillance systems. These systems are needed for monitoring patterns of environmentally driven pathogen/disease spillover, emergence/reemergence and spread (SES) and risk-based decision making and public health action.^11^

The United Nations Office for Disaster Risk Reduction (UNDRR) defines risk as the probability of an outcome having an adverse effect on people, systems, and/or assets.^12^ Risk analysis processes are utilized as an objective, systematic and comprehensive tool for the risk assessments of biological threats and to support evidence-based decision making by governments, international entities, and other key decision makers following an assessment of the risks posed by these evolving threats including those of zoonotic origins.^13^ Attempts have been made to integrate surveillance and risk modeling data on trends and projections about environmentally driven hazards and exposures as part of disaster risk management (DRM).^14^ However, insufficient attention has been given to the need for systematically incorporating findings about the increased threat posed by anthropogenic environmental changes to zoonotic biological threat SES into the entire spectrum of the risk analysis process (risk assessment, risk management and risk communication), to inform cross-sectoral pandemic prevention decision-making.^15^ Consequently, there remains limited understanding about the role of various environmental factors in the spillover and transmission dynamics of EIDs, including both arthropod-transmissible and non-arthropod transmissible EIDs with epidemic and pandemic potential. There is thus a need for the adoption of eco-epidemiological tools for the development of an integrated risk analysis (IRA) process due to the increasing evidence of environmentally driven EIDs in the East Africa region.^15^

### Aim & Objective

The aim of this integrative review was to fill gaps in knowledge and provide a holistic understanding about the environmental drivers of CCHFV, MARV, and MERS-CoV infections in the East Africa region to inform IRA processes at the environment-animal-human exposure interface and better integration of the environment sector into the operationalization of the One Health (OH) approach for risk analysis. The objective of the review was to utilize findings from the review for the initial conceptualization of an IRA framework. This framework adopts a combination of quantitative and qualitative data sources for risk analyses of environmentally driven EIDs using a OH approach.

The integrative review approach critically assesses and synthesizes evidence from both empirical and gray literature to develop a new conceptual or theoretical framework and perspective on an emerging research topic.^16^ An integrative review was regarded as the most appropriate method to utilize for our study considering the increasing recognition of the role of environmental drivers such as climate change as health security threat multipliers; and the need for systematically integrating these environmental drivers and other related cascading risk drivers under risk analyses to inform data-driven cross-sectoral decision making for pandemic prevention and biological risk reduction.

## METHODS

### Search strategy and record assessment

We conducted an integrative review search from June to December 2023, and an updated search in July 2024 using six bibliographic databases (Embase, PubMed, Web of Science, SCOPUS, CAB Direct, and CAB Digital) to identify peer-reviewed published literature. We also considered articles, book chapters, reports and other materials published in English and conducted a search for grey literature from the Africa Centers for Disease Control and Prevention (Africa CDC), Food and Agriculture Organization (FAO), WHO, World Organization for Animal Health (WOAH) and United Nations Office of Disaster Risk Reduction (UNDRR) websites. The search covered the period from 2000 to 2024, to account for the reported evidence of the increase in EIDs of zoonotic origins since the turn of the century.^1,2^ The three research questions underpinning the review included:

i. What are the environmental drivers of the spillover, emergence and/or reemergence, and spread of CCHFV, MARV, and MERS-CoV in the East Africa region?
ii. What risk frameworks and/or methodological tools exist for assessing and managing environmentally driven zoonotic EIDs and what are the gaps in existing risk analysis frameworks; and
iii. What types of policy recommendations (applicable to the East Africa region) are in place to address environmental drivers of emerging/reemerging zoonotic biological threats such as CCHFV, MARV, and MERS-CoV?

Studies and reports discussing environmental drivers of the three pathogens of focus (CCHFV, MARV, and MERS-CoV), and their respective diseases [Crimean-Congo haemorrhagic fever (CCHF), Marburg virus disease (MVD) and MERS] in the East Africa region were included in the study, The study also included risk frameworks/methodological tools, and policy recommendations to address the environmental drivers of zoonotic biological threats under risk analysis. For the review, we defined risk frameworks/methodological tools as decision-making frameworks, tools, and/or models that adopt structured processes and methods to assess, characterize, manage/mitigate, and communicate potential risks.

Search terms and strategies defining the pathogen/disease exposure, environmental drivers, risk framework/methodological tools, and policy questions were first developed and discussed extensively with a Library Informationist. A combination of relevant search terms was used for each question using keywords such as East Africa, Kenya, climate change, risk assessment, risk analysis, One Health, policy implementation, and environmental policy. A full list of the search terms utilized for the three review questions and inclusion criteria are included as an appendix [Supplemental Table 2 (ST2)].

Database articles search results were uploaded on Covidence (Veritas Health Innovation, Melbourne, Australia) according to the order of the three research questions guiding our overall study aim, to ensure rigor and maximum efficiency with data management, analysis, and synthesis.^17^Other types of relevant materials from the included websites were uploaded on the reference management software, Zotero (Corporation for Digital Scholarship, Fairfax, VA), for collation and inclusion of grey literature from online websites in the final list of selected articles.^18^

Adapting some components of the Preferred Reporting Items for Systematic Reviews and Meta-Analyses (PRISMA) 2020 expanded checklist,^19^ articles were screened via a two-step screening process based on title and abstract, and full text screening prior to final selection of articles by the main author for data extraction. Exclusion of articles were mainly due to wrong disease outcome and setting, no reference to a risk framework/methodological tool, non-environmental drivers (and/or a combination of the three), and non-English language. As further adapted from the PRISMA 2020 checklist, the method used for the process of selecting studies that met the study inclusion criteria was based on the relevance of the study or report to one and/or or a combination of the three research questions. A tagging system was used to track the search articles corresponding to the related research questions and main themes emerging from the review search.

### Critical appraisal of evidence

The data extraction process was carried out on Covidence using the data extraction template tool as a point of departure for the development of the final tool. Extracted data included study ID, Title, Authors, location, timeframe, environmental drivers, risk components, adopted risk framework/methodology, highlighted gaps, and policy recommendations. As part of the critical appraisal process, the final data extraction tool was piloted using 32 articles pre-identified as highly relevant across the three research questions guiding the search strategies (Supplemental Table 3 (ST3)]. The final sample of studies included a combination of both empirical studies (comprised different methodologies) and grey literature.

For the critical appraisal of evidence, we considered both the methodological rigor of the empirical studies and strength of the theoretical reports of the included articles and reports. In line with recommendations by Whittemore and Knafl (2005), we utilized a 2-point scale (high and low) to consider both the methodological or theoretical rigor of the retrieved empirical and grey literature and their level of relevance to the review questions.^20^ We utilized a thematic analysis for the synthesis of the evidence that corresponded to the three questions.

## RESULTS

A total of 18,075 studies published from 2000 to 2024 were identified and retrieved from the database searches, and 44 articles and reports identified via other sources (website searches). Of the total number of studies identified via database searches, 6611 were removed as duplicate records identified by COVIDENCE (n = 6484) and manually (n = 127). For the two-step screening process 11464 records underwent title and abstract screening out of which about 5% of these records (n = 521) were sought for retrieval for full text screening. The total number of records identified via database search and included in the review represented 43% of fully screened articles assessed for inclusion (n = 223). Following a similar step, 44 reports were identified via website searches and sought for retrieval, out of which 40 were assessed for inclusion and 48% (n = 19) were included in the review (*Figure 1*).

**Figure 1:**
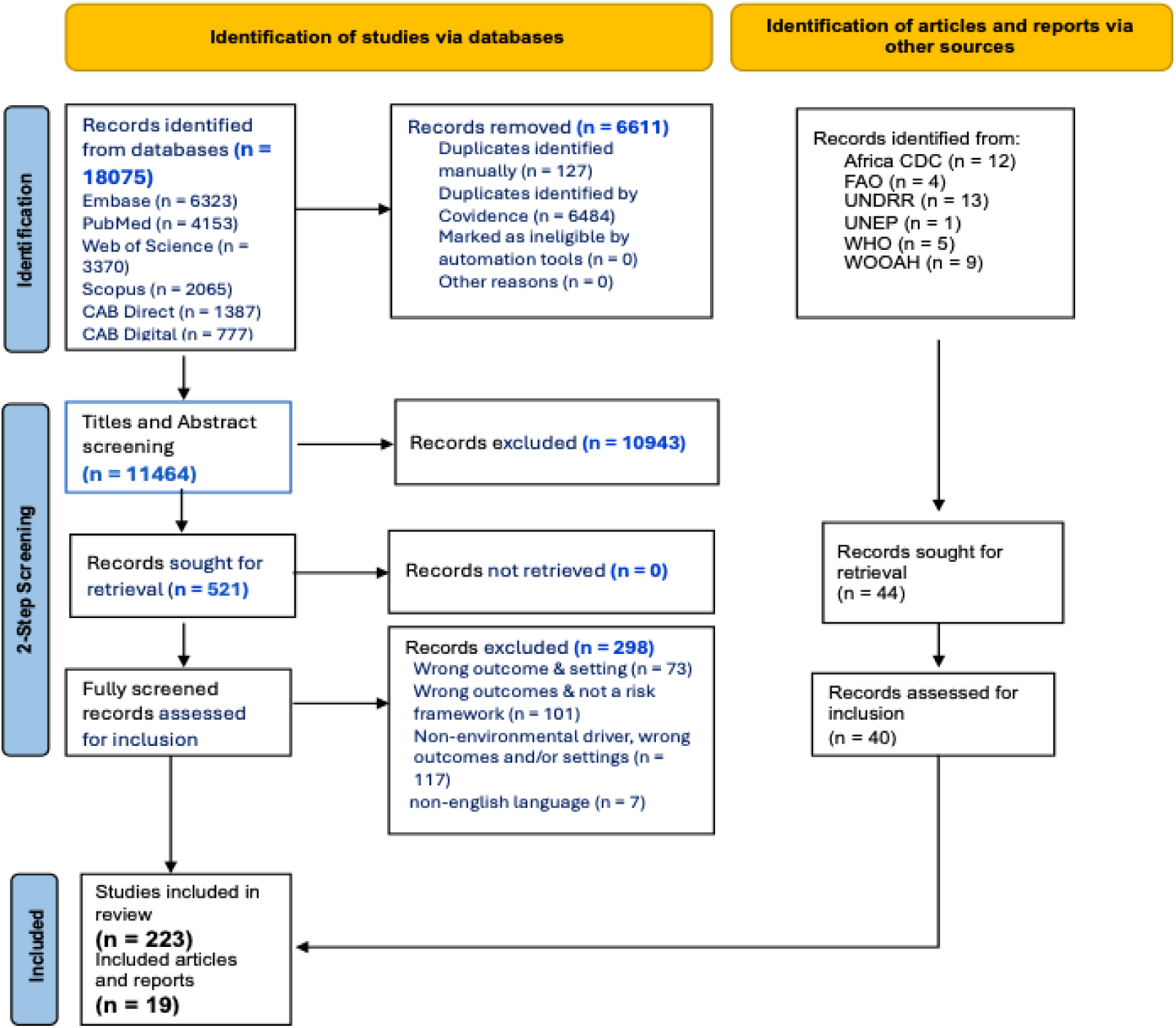
Article Screening and Selection Using the Preferred Reporting Items for Systematic Reviews and Meta-Analyses (PRISMA) Flow Diagram

### Characteristics of included studies and reports

Of the total number of retrieved studies and reports combined (n=242), the highest number of searches were relevant to the first review question on the environmental drivers (n=137), while the second highest search results were applicable to the second review questions on the risk frameworks and methodological tools (n=73), and the least number of searches covered the third review question related to policy recommendations (n=32), representing 57%, 30% and 13% of the total searches respectively (*Table 1)*.

**Table 1:** Search retrieval summary by review questions.

| **Review Question** | **Number of Searches Retrieved (database and website searches)** | **Percentage of Total number of included studies and reports** |
| --- | --- | --- |
| **Environmental Drivers** | 137 | 57% |
| **Risk Frameworks/Methodological Tools** | 73 | 30% |
| **Policy Recommendations** | 32 | 13% |
| **Total** | 242 | 100% |

In terms of geographic coverage, the most represented countries covered in the review included country-specific studies covering Kenya, Tanzania, and Uganda, and other countries in the region combined including Sudan, Ethiopia, and Madagascar. Other studies and reports covered multiple countries in East Africa, across the Sub-Saharan Africa region, and of global relevance inclusive of East African countries (*Figure 2*).

**Figure 2:**
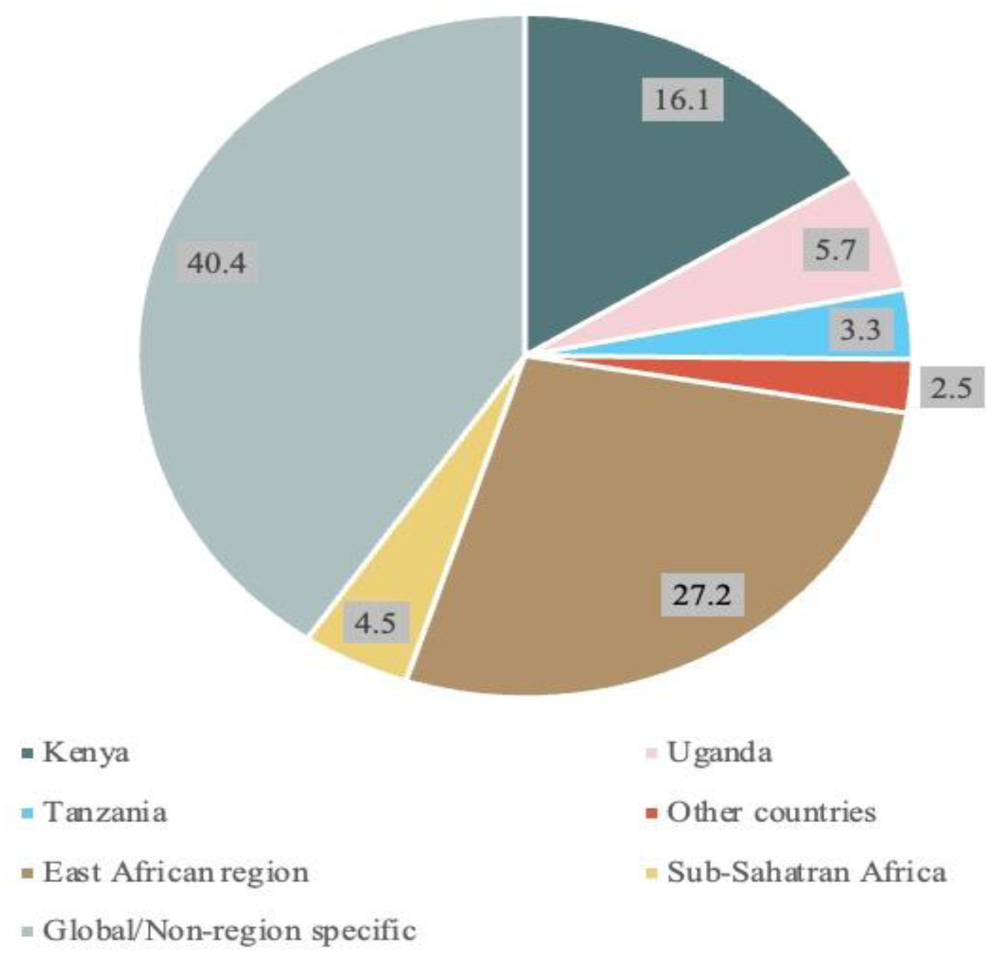
Percentage Distribution of Included Studies and Reports by Geographic Coverage

### Environmental Drivers

Our review findings highlighted 10 key environmental drivers of the spillover, emergence/reemergence and spread (SES) of CCHFV, MARV, and MERS-CoV in the East Africa region. These drivers include climate change, migration patterns, habitat encroachment, observed ecological changes, deforestation and reforestation, animal husbandry practices, livestock overgrazing, biodiversity loss, irrigation practices, and human-mediated transport of pathogens. An important theme emerging from our study finding was that nine of the ten identified environmental drivers all had a linkage to climate change, the tenth environmental driver and a cascading risk driver *(Figure 3*).^21^ For example, climate change was reported to have a direct impact on human, animal, and wildlife population migration patterns resulting in the geographic dispersal of CCHFV, MARV, and MERS-CoV to new areas in Ethiopia, Kenya, Sudan, and Uganda;^22–26^ human-mediated transport of pathogens such as MARV to new areas including from Uganda to neighboring Tanzania;^27^ migration of MARV reservoir hosts (cave-dwelling fruit bats) across Uganda for cultivated fruit farming foraging activities;^28^ irrigation practices that provide additional breeding sites for CCHFV-infected ticks and pathogen emergence into new areas;^29^ and biodiversity loss.^30^

**Figure 3:**
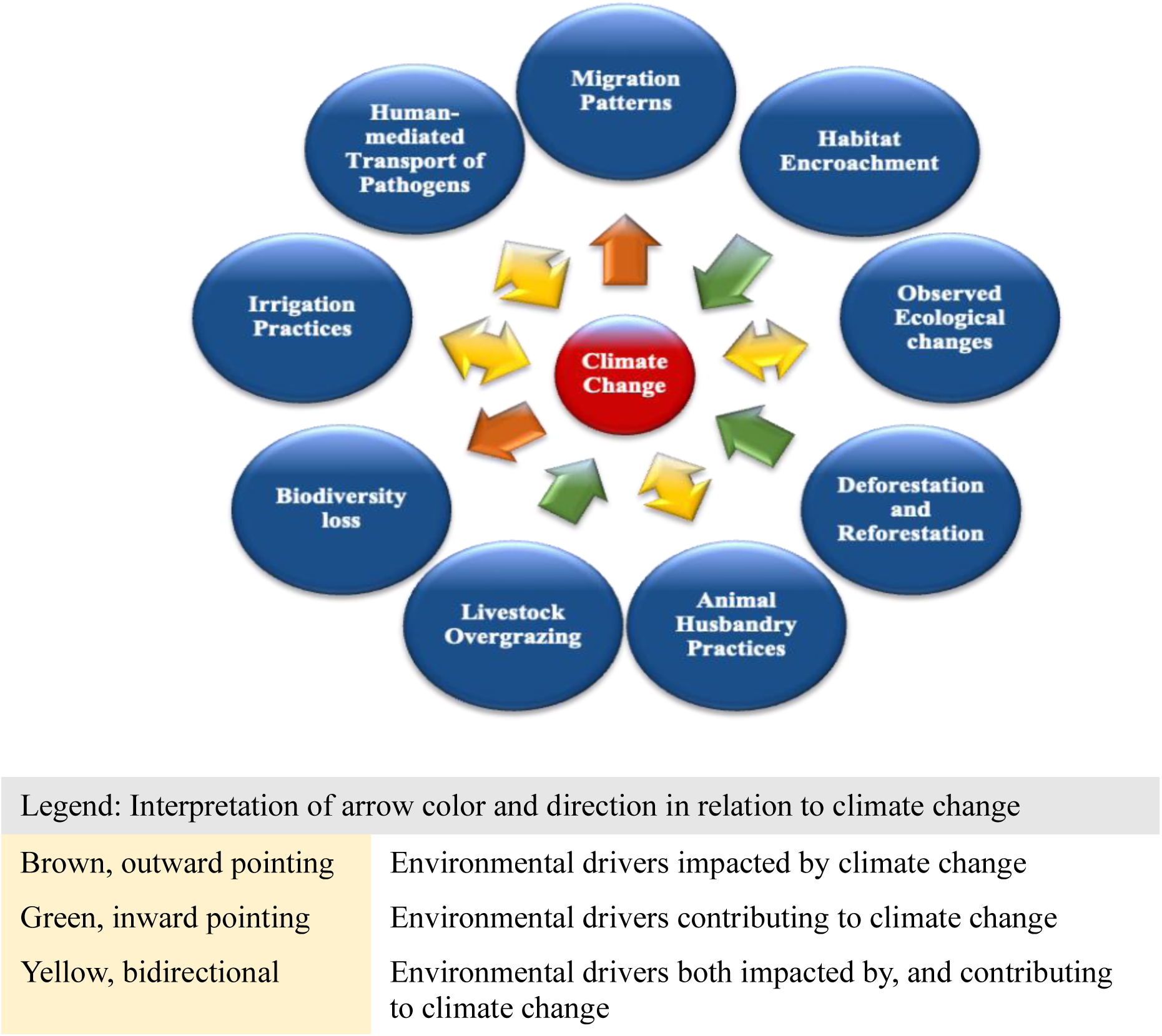
Environmental Drivers of Emerging/Reemerging Infectious Diseases in the East Africa Region

Other environmental drivers were reported to contribute to climate change including via the emission of greenhouse gases (GHGs), as we found for habitat encroachment linked to mining activities and multiple outbreaks of MARV in Uganda;^22,31,32^ observed ecological changes associated with CCHFV spillover events;^33^ deforestation and reforestation associated with CCHF, MVD, and MERS transmission across countries in the region;^34,35^ animal husbandry practices associated with an increased risk for MERS-CoV transmission,^36,37^ and livestock overgrazing linked to MVD and CCHF transmission in Kenya and Uganda.^38–40^ Reported environmental drivers like animal husbandry practices, human mediated transport of pathogens, irrigation practices, and observed ecological changes were observed to have a bidirectional relationship in their linkage to climate change.

In supplemental table 4 (ST4), we have presented the direct and indirect impact of climate change associated with other identified environmental drivers and pathogen transmission routes, using examples of evidence (epidemiological, ecological and/or serological) from the literature.

### Risk frameworks and methodological tools for assessing environmental drivers of zoonotic biological threats

Several studies highlighted risk-based approaches and methodological tools useful to determine the geographic distribution of infectious disease pathogens in relation to pathogen/disease SES in the East Africa region (and their application across other regions, where applicable). Some included studies also discussed risk analysis data gaps and the need for environmental, climatological, socio-anthropogenic, and socio-economic variables for driving risk predictions and characterization as part of IRA.^23^

Findings on existing risk frameworks and methodological tools are categorized under four key themes:

i. *Risk mapping-based assessment tools:* Several studies discussed the application of both conventional and specialized risk mapping approaches as part of risk assessments of zoonotic biological threats including the integration of the FAO Rift Valley fever decision support tool (RVF-DST) risk mapping data with geospatial and surveillance data to enhance early warning and forecasting capacities.^41^ A reported limitation with conventional risk maps is the limited capacity to properly characterize risks and risk levels in areas where there are data gaps due to low surveillance coverage.^42^ As a specialized risk mapping approach, Ecological Niche Modeling (ENM), also referred to as Species Distribution Modeling (SDM), is used to fill surveillance data gaps, including for determining CCHF prevalence and geographic distribution of ticks in Sudan,^23^ to improve surveillance of CCHF hotspots in Kenya, South Sudan, and Tanzania,^43^ and to determine environmental and socio-ecological suitability for CCHF infections in Uganda;^33^ utilizes environmental data [such as enhanced vegetation index (EVI), normalized difference vegetation index (NDVI), land surface temperature, and potential evapotranspiration] to enable real-time monitoring of vulnerable populations and potential disease transmission pathways including identifying hotspots for MVD transmission across the Africa region and populations at risk of MARV spillover.^44^ We compiled a list of tested predictor variables utilized for ENM and SDM and reported to have a significant association with risk of CCHFV, MARV, and MERS-CoV emergence, transmission, and spread (*Table 2*).
ii. *Multivariable risk assessment and management frameworks*: incorporating a combination of different categories of variables for multihazard risk assessment including the Index For Risk Management Epidemic Risk Index (INFORM ERI) adapted from the INFORM GRI;^46,47^ Spatial Multicriteria Decision Analysis tool ^48^ WHO Health Emergency and Disaster Risk Management (EDRM) framework;^49^ UNDRR Technical guidance framework;^50^ and the integrated risk and vulnerability assessment framework.^51^
iii. *Qualitative and semi-quantitative risk tools based on expert opinions*: Some studies reported the use of expert-informed qualitative risk assessment tools such as the Tripartite Joint Risk Assessment Operational Tool (JRA OT).^52^ Other risk assessment tools informed by expert opinions include Spillover Viral Risk Ranking tool;^53^ infectious disease seeker software tool;^54^ and a semi-quantitative risk assessment tool utilized for the risk assessment of spillover routes and disease amplification.^55^
iv. *Quantitative-based environmental risk assessment tools: i*ncluding Quantitative Microbial Risk Assessment;^56^ and Bayesian Belief Networks ^57^

**Table 2:** Summary of tested predictor variables with high predictive accuracy for risk of CCHFV, MARV, and MERS-CoV emergence, transmission, and spread.

| Variable Category | CCHFV | MARV | MERS-CoV |
| --- | --- | --- | --- |
| **Environmental/Socio-environmental** | ▪ Landcover<br>▪ Average Annual Change in Nighttime Light Index (measures shift in population density/increase in development and human population size).<sup>45</sup><br>▪ Cattle Density<br>▪ Shrub type<br>▪ Enhanced Vegetation Index<br>▪ Normalized Difference Vegetation Index<br>▪ Percent sand-soil content | ▪ Enhanced Vegetation Index<br>▪ Elevation level | ▪ Population density (human and camels)<br>▪ Landcover<br>▪ Bare land coverage<br>▪ Forest coverage<br>▪ Dry season grazing area |
| **Climate** | ▪ Land Surface Temperature (mean temperature of wettest and driest quarter of the year)<br>▪ Precipitation (of wettest quarter of the year) | ▪ Temperature seasonality<br>▪ Rainfall (mean annual, driest quarter) | ▪ Annual Mean Temperature<br>▪ Frequency of Drought events<br>▪ Humidity<br>▪ Wind Speed |

As a summary, we have presented key aspects of the risk frameworks/methodological tools covered in the review including unique features and benefits, areas of application, extent of geographic coverage, and highlighted gaps and areas for improvement [Supplemental Note 1 and Table 5 (ST5)]. Our findings highlight the need to properly account for environmental risk drivers of pathogen/disease SES by using a combination of quantitative and qualitative data sources to improve risk characterization and to better inform OH risk mitigation and communication interventions.

### One Health policy recommendations for risk analysis

A few articles and reports discussed policy recommendations to improve IRA processes by enhancing the integration of the environment sector into the OH approach. These recommendations fall under three main categories:

i. *Incorporating pathogen risk assessment under environmental assessments/management plans*: Some articles and reports identified the need to enforce the conduct of zoonotic spillover risk assessments as part of environmental impact assessments and land-use planning;^58–62^ account for infectious diseases risk in environmental impact and risk assessments as a pandemic prevention and preparedness strategy;^61,63–65^ and include zoonotic spillover risk reduction measures in management plans of high-risk mining, logging, ranching (livestock keeping), and infrastructure programs.^2^
ii. *Addressing data gaps for better risk characterization*: Recommendations for addressing data gaps to improve risk characterization include application of socioecological models to characterize disease emergence risk;^66,67^ and use of remote sensing data on land use practices, climate change and other environmental variables, population vulnerabilities, and socio-economic indicators; identified studies recommend combining these data with epidemiologic data for the development of risk maps as part of EID SES risk analysis to inform decision making and the implementation of interventions targeted at vulnerable populations.^68–71^
iii. *Implementation of data-driven risk mitigation interventions*: A recent scoping review focused on the East Africa region discussed recommendations from different studies on the need to develop and implement environmental monitoring policies as a key component of risk analysis.^30^ Highlighted policy interventions include routine monitoring of wastewater from abattoirs for presence of EIDs pathogens and other pathogen types using shotgun metagenomics sequencing (for enhancing surveillance capacities);^30^ improving biosecurity and establishing a biosurveillance network in the region;^30^ routine use of climate data and spatial techniques to provide early warning about EIDs and other biological threats;^72,73^ linkage of climate change mitigation strategies to OH programs;^74–76^ and systematic integration of earth observation and geospatial data into national risk reduction decision structures.^77^

Another study discussed measures that have been taken to address climate change and health policy gaps in the region to increase awareness and to inform decision and accountability mechanisms;^78^ and the Enhancing National Climate Services (ENACTS) initiative focused on improving the availability, access, and use of climate information for decision making across health and other sectors.^79^ Other policy recommendations include integration of climate change adaptation and mitigation policies into health policies as part of risk analysis to reduce population vulnerabilities to EIDs risks;^80,81^ and improving weather forecasting including forecasting of EWEs and meteorological hazards and development of regionally sustainable EWS on climate change predictions known to affect disease transmission.^35^

Our findings across the three review questions highlighted the need to adopt an IRA framework that combines quantitative data on known environmental drivers of EIDs (obtained via specialized risk mapping-based techniques like ENM and remote sensing data) with socioanthropogenic and qualitative data informed by expert opinions as part of risk assessment and risk characterization processes (*Figure 4*). We conceptualized the IRA framework using the three dimensions of risk defined under the INFORM GRI (hazard & exposure, population vulnerability, and lack of coping capacity).^14,46^ The framework comprises a 4-tiered process with the first 3 stages (integrated risk assessment, zoonotic biological threat risk characterization, and cross-sectoral risk management and risk communication) in line with basic concepts of risk analysis. Our findings also support the adoption of risk mitigation and communications interventions under the framework, as adapted from the WHO Health EDRM framework, and via the implementation of cross-sectoral policies to address lack of coping capacities that increases population vulnerabilities to EID risks. The IRA framework also includes a fourth evaluation stage that captures essential factors to consider in evaluating the level of enhanced coping capacities due to the three preceding IRA stages. Inclusion of this evaluative component under the framework is in accordance with identified policy recommendations for improving risk governance and regulations.^82–84^

**Figure 4:**
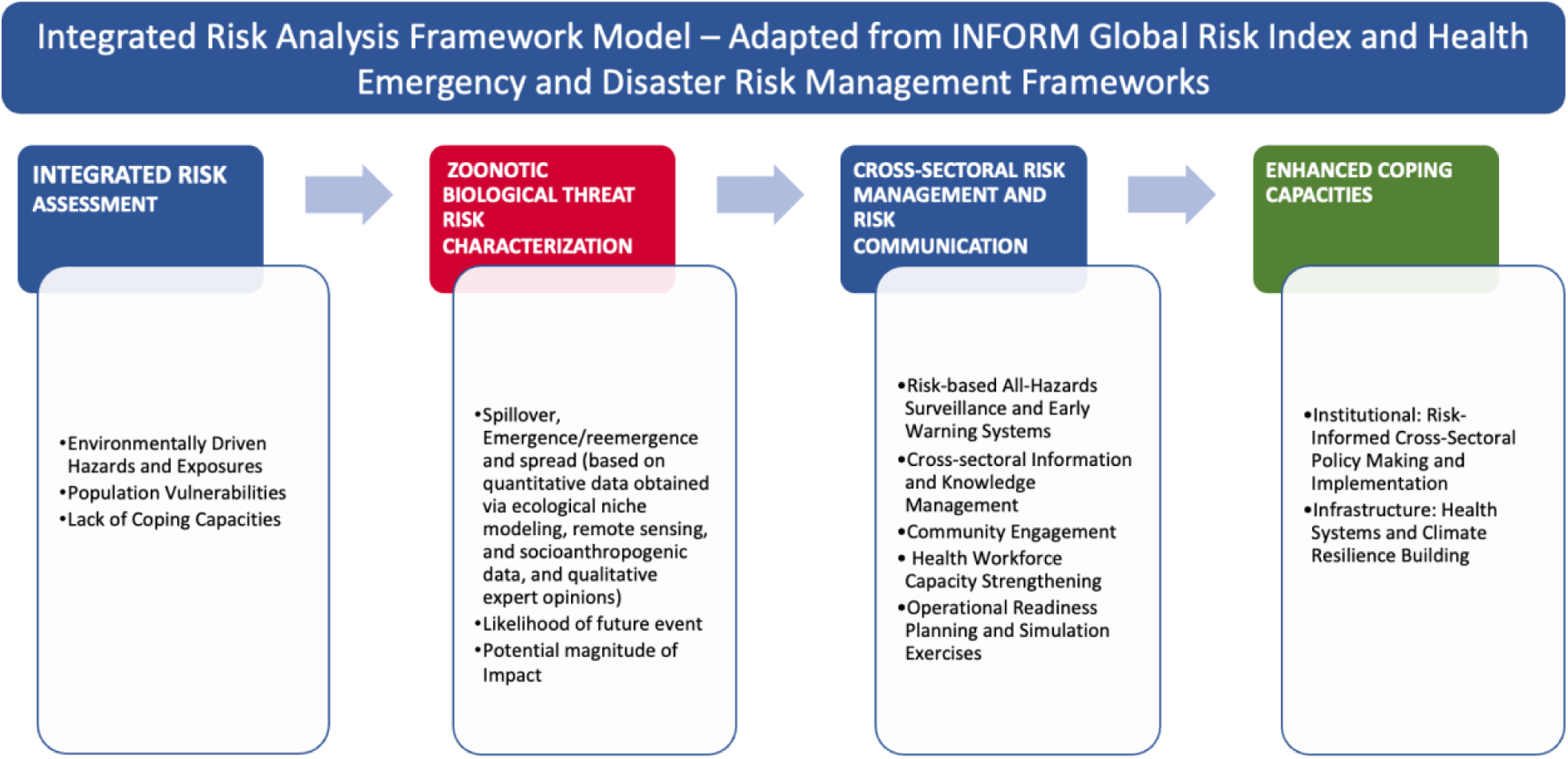
Integrated Risk Analysis conceptual framework model incorporating ecological niche modeling, remote sensing data and qualitative expert opinions (findings from integrative review)

## DISCUSSION

This review contributes to identification of knowledge gaps and provision of a holistic understanding about the impact of environmental drivers like climate change on the SES of zoonotic EIDs of epidemic and pandemic potential in the East Africa region. Our review findings highlight how climate change serves as a cascading risk driver associated with risks of outbreaks of viral EIDs like CCHF, MVD, and MERS. These findings are in line with a previous study describing how the effects of climate change on cross-species viral transmission risk could likely cascade in the future emergence of viral zoonoses.^85^ With increasing cases of CCHF and MVD outbreaks and evidence of CCHF and MERS seroprevalence reported across countries in the East Africa region,^86–89^ there is a need for data-driven and risk-based decision making for prevention of pandemics and other severe outbreaks that could pose a GCBR. In line with our review findings, the UNDRR describes climate change as an amplifier of biological risks with escalation potential worsened by population vulnerabilities and lack of coping capacities.^21^

### Climate Change as a cascading risk driver

Using a causal loop diagram to interpret our study findings on the environmental drivers, we describe both the direct and indirect impacts of climate change and cascading risk pathways that could potentially increase the risk of a severe pandemic due to an uncontained outbreak of CCHF, MVD, and/or MERS (*Figure 5*). The diagram highlights three primary anthropogenic reinforcing loops that serve as amplifiers of SES of these three EIDs and their causative pathogens and may contribute to an increase in the risk of an outbreak of catastrophic magnitude such as a severe pandemic.

**Figure 5:**
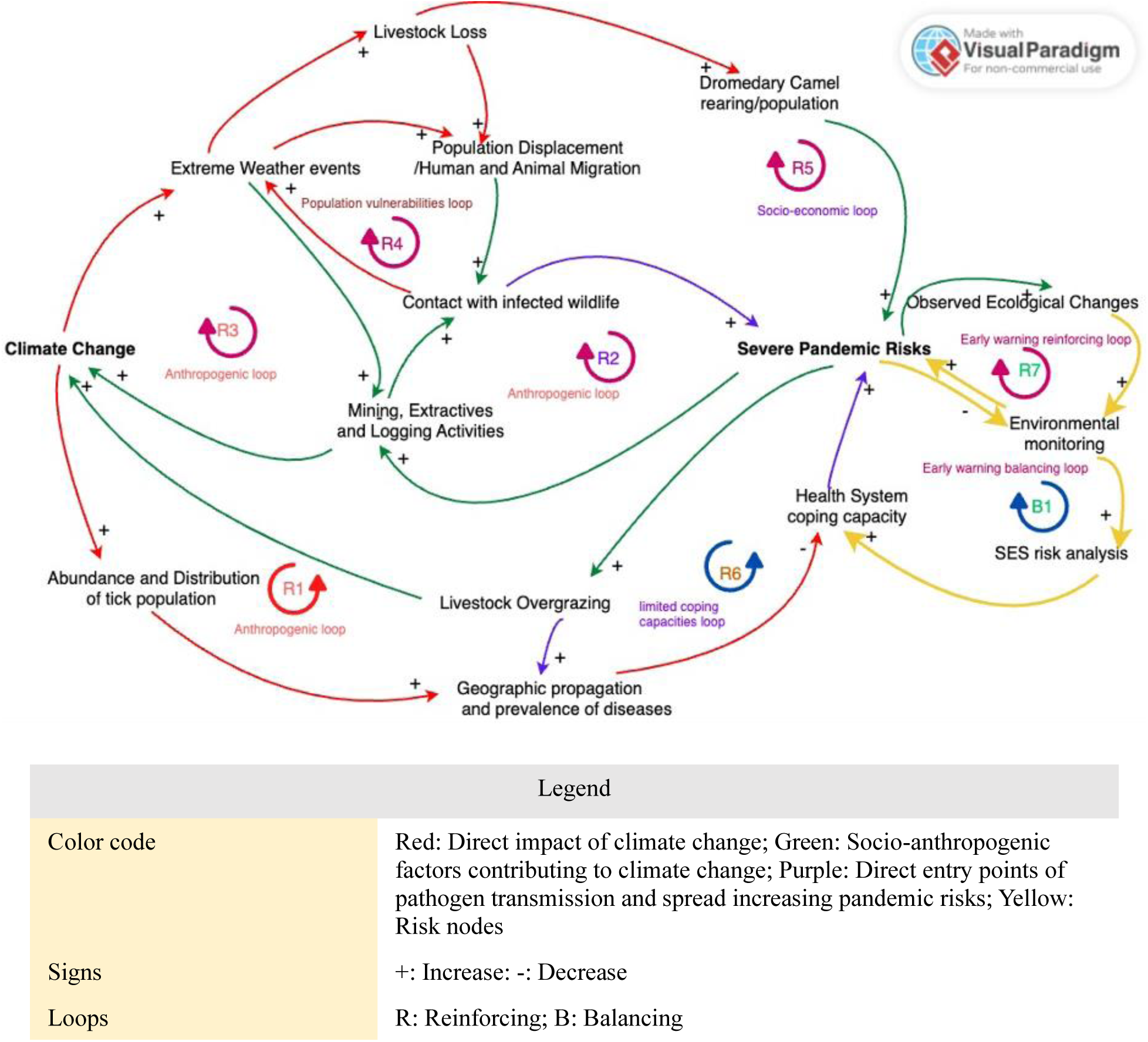
Causal Loop Diagram of the direct and indirect impact of climate change and cascading risk pathways contributing to severe pandemic risks

The first anthropogenic reinforcing loop relates to the impact of climate change on the increasing abundance and distribution of disease-infected tick population and a consequent increase in the geographic propagation and prevalence of tick-borne diseases like CCHF.^90,91^ Along with limited coping capacity of health systems, the risk of a severe pandemic is further amplified by socio-anthropogenic risk drivers of CCHFV transmission such as livestock overgrazing, which further contributes to climate change.^24,38^

For the second anthropogenic loop, habitat encroachment related to mining, extractives and logging activities increases human contact with MVD-infected reservoir host species (fruit bats) roosting along the periphery of their natural habitat (caves and mines).^32,92^ These activities further contribute to GHG emissions and climate change impact that increase severe pandemic risk.^93^

The impact of activities that contribute to GHG emissions also relates to the third anthropogenic loop by which EWEs due to climate change result in livestock loss and population displacement, in line with our review findings.^29^ A combination of the anthropogenic amplifiers, population vulnerabilities, and socioeconomic reinforcing loops may increase the risk of a severe pandemic due to an uncontained outbreak of MERS. For this risk pathway, climate change-induced population displacement and migration increases contact with infected wildlife, while increase in dromedary camel population and adoption of camel rearing as a form of drought resilience increases the risk of MERS-CoV spillover to humans and a consequent MERS outbreak.^25,86^

The inclusion of an early warning loop outside of the main cascading risk causal loop system reinforces the need for a SES risk analysis as a strategy for introducing balance to the system by identifying risk nodes and critical control intervention points for targeting pandemic risk reduction efforts and future spillover prevention measures.

### Utilizing Ecological Niche Modeling to fill data gaps in risk analysis processes

Increasing evidence of climate change induced biological risk drivers in the region and identified gaps in existing risk frameworks and methodological approaches warrant the application of environmental monitoring-based early warning tools like ENM, to improve our understanding of the risk pathways of environmentally driven EIDs. In line with previous study findings, these specialized risk mapping techniques are required as an integral part of risk analysis for the development and implementation of appropriate data-driven pandemic prevention and biological risk mitigation policies and strategies.^94^ Although ENM and SDM were used interchangeably in some studies, the distinct feature between the two terms is in reference to the mapping of potential distribution of species for ENM and actual species distribution within a geographic coverage for SDM.

Findings from our review highlight the need for utilizing data driven risk analysis processes to inform the implementation of future cross-species pathogen spillover and EID outbreak prevention strategies. Our findings further emphasize how limited data on environmentally driven spillover transmission risk contribute to inaccuracies in risk characterization and subsequent gaps in risk management and communication strategies. These findings are in line with findings from the study by Jones et al. (2017) about gaps in traditional risk analysis approach to disease management; the study highlights the need to incorporate ecological and socio-economic drivers of disease risk into an integrated eco-social risk analysis framework for analyzing risk of infectious disease emergence and spread.^67,95^ Sorvillo et al. (2020) discussed the need for joint risk assessments of zoonotic biological threats like CCHFV by utilizing a comprehensive framework that combines epidemiological, ecological, virology and vector biology data, with modeling data to make an estimation of outbreak risk and target surveillance activities.^96^ Our review findings also align with recent calls for adopting model-based risk assessments to assess current biological threats and to predict future risks of zoonoses emergence and/or reemergence linked to environmental factors such as climate change to target surveillance needs, and to enhance overall pandemic prevention, preparedness, and response efforts.^97,98^

### Implications for research and public health practice

Our review findings were used to inform the initial development of a newly conceptualized IRA framework that can be adopted at both national and sub-national levels for framing the conduct of environmentally driven zoonotic biological threat risk assessments using both quantitative and qualitative data sources. The framework is helpful for better characterizing the risk of SES of environmentally driven EIDs to inform the adoption and implementation of evidence-informed risk management and communication interventions, and OH strategies, policies, and action plans. With competing priorities identified as a primary barrier to cross-sectoral collaboration for disease surveillance,^99^ there is an opportunity to utilize a risk-based model under the IRA framework to inform efficient resource allocation. The proposed framework is in alignment with the work of the One Health High Level Expert Panel (OHHLEP) on reducing the risk of zoonotic spillovers at the source via improved prevention approaches;^100^ the preventing zoonotic disease emergence (PREZODE) initiative that aims to provide support to countries to adopt a risk analysis-based approach to prevent the emergence and spread of zoonotic diseases;^101^ and the World Bank OH holistic investment framework to mitigate cascading pandemic risks and their impacts on national, regional, and global economies.^102^

Based on the preliminary background review and the integrative review findings, the IRA framework can be operationalized using the example of climate change induced CCHF cases in Kenya [Supplemental Note 1 (SN1)]. The figure in SN1 describes how the IRA conceptual framework model could be applied to provide a coordinated cross-sectoral response to a hypothetical future cluster of CCHF cases in Kenya. Overall, components of the newly conceptualized IRA framework represent a blueprint for enhancing the integration of the environment sector into the operationalization of the OH approach at the national and sub-national level, via the proper integration of environment sectoral policies, strategies, and interventions. Adopting an integrated approach will contribute towards a more effective cross-sectoral response to EIDs outbreaks of zoonotic origins and proactive preparedness planning to address future health security threats.

### Limitations

There are a few study limitations to keep in mind while interpreting the study findings. First, most of the study findings on risk methodology to improve our understanding about the environmental drivers of zoonotic EIDs are largely model based. Thus, there is a need for further empirical research and validation across all relevant sectors in consultation with key stakeholders including technical experts and policy makers. Validation of integrated risk models would require close cross-sectoral collaboration including among different areas of expertise such as climate, health, earth observation, and geospatial experts to reach a consensus on important variables that would be the best fit for risk assessment and prediction models. Utilization of these variables are needed to inform allocation of resources such as for surveillance activities, pandemic prevention, and OH policy development. Further, relevant studies referenced in our review also highlight the need for overcoming inherent bias and uncertainties in available data used for making risk predictions.^103^

Although not a requirement under integrative review studies, the screening and selection process could have benefited from having multiple people screen the individual studies and reports included in the review. To strengthen the process, we utilized the recommended approach for critical appraisal of evidence under integrative reviews.^20^

## CONCLUSION

Increasing cases of the spillover, emergence and/or reemergence and spread of zoonotic EIDs of pandemic potential like CCHF, MVD, and MERS have been attributed to environmental drivers all linked to climate change. The potential threat and impact of climate change as a cascading risk driver calls for adopting a model-based and data-driven IRA approach across regional, national, and sub-national levels to better characterize the risks posed by environmentally driven EIDs in the East Africa region. Improved risk characterization is needed to make more accurate projections about potential geographic distributions of EIDs to new areas.

Based on our review findings, we recommend the adoption of ENM and spatial analytic techniques like geospatial analysis and remote sensing tools to develop risk maps for risk analysis of the environmental drivers of EIDs SES. These specialized risk mapping tools serve as early warning systems to inform resource allocation, decision making and policy implementation using a OH approach. ENM-based risk mapping also holds promise to inform primary preventions efforts focused on the implementation of ecological countermeasures against environmental drivers of cross-species pathogen spillover and EID outbreaks.

Our study findings highlight the need for including tested environmental variables like NDVI, EVI, proportion of forested land, and non-irrigated agricultural land cover; and climate change variables such as annual mean precipitation, land surface temperature, and humidity levels to develop regional, national, and sub-national/local disease risk maps using ENM and remote sensing tools, especially in areas with limited surveillance and reporting capacities. Utilizing a combination of ENM data and other quantitative and qualitative data sources under an IRA framework model will also help enhance the rigor and quality of the overall risk analysis process including to make further assessments about the pandemic potential of EID pathogens.

Considering the increasing risk of distribution and propagation of environmentally driven EIDs to new areas, data-driven IRA processes will help inform climate-resilient health systems building and enhancement measures and decision making for pandemic prevention. Overall, our findings call for promoting practical cross-sectoral climate change mitigation policies under IRA processes as central to pandemic prevention and GCBR reduction.

## Data Availability

No external dataset were used under this study. All data produced for the study are contained in the manuscript.

## Author contribution

All authors contributed to the design and drafting of the final manuscript. OA led the study conception and design, and the writing and finalization of the manuscript. LR contributed to the methodology section. JN and MD led the supervision of the study. CW led the study funds acquisition process. All authors agreed to submit the final manuscript for publication.

## Ethics Statement

This study does not involve human participants

