## Supplemental Materials for "Improving risk analysis of the environmental drivers of the spillover, emergence/reemergence, and spread of Crimean-Congo haemorrhagic fever virus, Marburg virus, and Middle East respiratory syndrome coronavirus in the East Africa Region"

*Supplemental Table 1 (ST1): Summary of reported outbreaks of Crimean-Congo haemorrhagic fever (CCHF) and Marburg virus disease (MVD) and evidence of Crimean-Congo haemorrhagic fever virus (CCHFV) and Middle East respiratory syndrome coronavirus (MERS-CoV) seroprevalence across countries in the East Africa region*

| Pathogen | Disease | Year of Outbreak | Country of origin | Source of Transmission |
|---|---|---|---|---|
| **CCHFV** | CCHF | 2022[1] | Uganda | Human |
| | | 2018-2019[2] | Uganda | Human |
| | | 2018[3] | Uganda | Human |
| | | 2013 - 2017[3] | Uganda | Human |
| | | 2015 - 2016[4,5] | Sudan | Nosocomial and Human |
| | | 2008 - 2009[6] | Madagascar | Tick and Human |
| | | 2009[7] | Sudan | Nosocomial and Human |
| | | 2008[7] | Sudan | Nosocomial and Human |
| | | 2000[8] | Kenya | Tick and Human |
| **MARV** | MVD | 2024[9] | Rwanda | Cave visit |
| | | 2023[10] | Tanzania | Inconclusive |
| | | 2007[11] | Uganda | Kitaka Mines |
| | | 2017[12] | Uganda | Mines and cave visits |
| | | 2014[13] | Uganda | Mines and cave visits |
| | | 2012[14,15] | Uganda | Mine and cave visits |
| | | 2008 (July)[16] | Uganda | Gold mine and cave visits |
| | | 2008 (January)[17] | Uganda | Cave visits |
| **Pathogen** | **Disease** | **Year** | **Location** | **Evidence of Seroprevalence** |
| **CCHFV** | CCHFV | 2024[18] | Northern Tanzania | 15.1-49.6% |
| | CCHFV | 2021[19] | Kenya | |
| | CCHFV | January 2020 – December 2021[20] | Kenya | 11.9 % seroprevalence among livestock [donkeys [31.4%), cattle (14.1%), sheep (9.8%), goats (8.1%)] |
| | CCHFV | 2012[21] | Kenya | |
| | MERS | 2018 – 2020[22,23] | Kenya | 0.2 % seroprevalence (3 humans tested positive); 100% for camels |
| **MERS-CoV** | | January 2016 – June 2018[24] | Across Kenya | 82% seroprevalence among adult camels |
| | | January 2016 – June 2018[22,24] | All (13) camel-rearing counties in Kenya | 68% seroprevalence across all counties combined |

| | | | | |
|---|---|---|---|---|
| | | 2016-2018[25] | Northeast Region, Uganda | 66% and 65% seroprevalence in camels from Morto and Amudat respectively |
| | | 2017[26] | Somalia | 100% seroprevalence in camel sera |
| | | 2017[26] | Sudan | 100% seroprevalence in camels |
| | | October – November 2016[27] | Kenya | 8.6% seroprevalence among camel herders |
| | | 2016[28] | Ethiopia | 70% seroprevalence in camels |
| | | 2015[26] | Sudan | 92% seroprevalence in camels |
| | | 2013 - 2014[29] | Garissa and Tana River counties, Kenya | Evidence of seropositivity in humans |
| | | 2013[30,31] | Marsabit county, Kenya | 90% seroprevalence among camels |
| | | 2010 - 2011[32] | Ethiopia | 93% and 97% seropositivity reported in Juvenile and adult camels respectively |

*Supplemental Table 2 (ST2): List of review search terms and inclusion criteria by questions*

| Review Question | Database Search terms |
|---|---|
| **1. What are the environmental drivers of the spillover, emergence/reemergence, and spread of Crimean-Congo haemorrhagic fefer virus (CCHFV), Marburg virus (MARV), and Middle East respiratory syndrome coronavirus (MERS-CoV) in the East Africa region?**<br><br>Inclusion criteria<br>- Environmental drivers<br>- Outcome of Interest (CCHFV/Crimean-Congo haemorrhagic fever (CCHF), MARV/Marburg virus disease (MVD), MERS-CoV/Middle East respiratory syndrome (MERS)<br>- East Africa<br>- English Language | **Pubmed**<br>"Africa, Eastern"[Mesh] OR "Africa east"[tiab:~2] OR "Africa eastern"[tiab:~2] OR "African east"[tiab:~2] OR "African eastern"[tiab:~2] OR "africans east"[tiab:~2] OR "africans eastern"[tiab:~2] OR Burundi*[tw] OR Comoros*[tw] OR Djibouti*[tw] OR Eritrea*[tw] OR Ethiopia*[tw] OR Kenya*[tw] OR Madagascar*[tw] OR Mauritius* [tw] OR Rwanda*[tw] OR Seychelles*[tw] OR Somalia*[tw] OR Sudan*[tw] OR Tanzania*[tw] OR Uganda*[tw] OR "Addis Ababa"[tw] OR Antananarivo[tw] OR Asmara[tw] OR Bujumbura[tw] OR "Dar es Salaam"[tw] OR "Dire Dawa"[tw] OR "Djibouti City"[tw] OR Dodoma[tw] OR Gitega[tw] OR Hargeisa[tw] OR Juba[tw] OR Kampala[tw] OR Keren[tw] OR Khartoum[tw] OR Kigali[tw] OR Kisumu[tw] OR Lindi[tw] OR Mahajanga[tw] OR Mbarara[tw] OR Mogadishu[tw] OR Mombasa[tw] OR Muhanga[tw] OR Moroni[tw] OR Muyinga[tw] OR Mwanza[tw] OR Nairobi[tw] OR Ngozi[tw] OR Omdurman[tw] OR "Port Louis"[tw] OR Victoria[tw] OR "Great Lakes Region*"[tw] OR "Horn of Africa*"[tw] OR "Nile Valle*"[tw]<br>AND<br>"Hemorrhagic Fever Virus, Crimean-Congo"[Mesh] OR "Crimean-congo hemorrhagic fever*"[tw] OR "Crimean-Congo haemorrhagic fever*"[tw] OR CCHF[tw] OR "congo virus*"[tw] OR "Crimean hemorrhagic fever*"[tw] OR "Marburg Virus Disease"[Mesh] OR Marburg[tw] OR MVD[tw] OR "Middle East Respiratory Syndrome Coronavirus"[Mesh] OR "Middle East Respiratory syndrome coronavirus"[tw] OR "MERS-CoV"[tw] OR "MERS Virus*"[tw] OR "Middle East respiratory syndrome"[tw] OR Merbecovirus*[tw]<br><br>**Embase**<br>'east african'/exp OR ((Africa* NEAR/2 east*) OR Burundi* OR Comoros* OR Djibouti* OR Eritrea* OR Ethiopia* OR Kenya* OR Madagascar* OR Mauritius* OR Rwanda* OR Seychelles* OR Somalia* OR Sudan* OR Tanzania* OR Uganda* OR "Addis Ababa" OR Antananarivo OR Asmara OR Bujumbura OR "Dar es Salaam" OR "Dire Dawa" OR "Djibouti City" OR Dodoma OR Gitega OR Hargeisa OR Juba OR Kampala OR Keren OR Khartoum OR Kigali OR Kisumu OR Lindi OR Mahajanga OR Mbarara OR |

| | |
|---|---|
| | Mogadishu OR Mombasa OR Muhanga OR Moroni OR Muyinga OR Mwanza OR Nairobi OR Ngozi OR Omdurman OR “Port Louis” OR Victoria OR “Great Lakes Region*” OR “Horn of Africa*” OR “Nile Valle*”):ab,ti,kw<br>AND<br>'Crimean-Congo hemorrhagic fever virus'/exp OR 'Marburg hemorrhagic fever'/exp OR 'Middle East respiratory syndrome coronavirus'/exp OR ("Crimean-congo hemorrhagic fever*" OR "Crimean-Congo haemorrhagic fever*" OR CCHF OR "congo virus*" OR "Crimean hemorrhagic fever*" OR Marburg OR MVD OR "Middle East Respiratory syndrome coronavirus" OR "MERS-CoV" OR "MERS Virus*" OR "Middle East respiratory syndrome" OR Merbecovirus*):ab,ti,kw<br><br>**Scopus**<br>TITLE-ABS-KEY ((Africa* W/2 east*) OR Burundi* OR Comoros* OR Djibouti* OR Eritrea* OR Ethiopia* OR Kenya* OR Madagascar* OR Mauritius* OR Rwanda* OR Seychelles* OR Somalia* OR Sudan* OR Tanzania* OR Uganda* OR "Addis Ababa" OR Antananarivo OR Asmara OR Bujumbura OR "Dar es Salaam" OR "Dire Dawa" OR "Djibouti City" OR Dodoma OR Gitega OR Hargeisa OR Juba OR Kampala OR Keren OR Khartoum OR Kigali OR Kisumu OR Lindi OR Mahajanga OR Mbarara OR Mogadishu OR Mombasa OR Muhanga OR Moroni OR Muyinga OR Mwanza OR Nairobi OR Ngozi OR Omdurman OR "Port Louis" OR Victoria OR "Great Lakes Region*" OR "Horn of Africa*" OR "Nile Valle*")<br>AND<br>TITLE-ABS-KEY ("Crimean-congo hemorrhagic fever*" OR CCHF OR "congo virus*" OR "Crimean hemorrhagic fever*" OR "Crimean-Congo haemorrhagic fever*" OR Marburg OR MVD OR "Middle East Respiratory syndrome coronavirus" OR "MERS-CoV" OR "MERS Virus*" OR "Middle East respiratory syndrome" OR Merbecovirus*)<br><br>**Web of Science**<br>TS=("Crimean-congo hemorrhagic fever*" OR CCHF OR "congo virus*" OR "Crimean hemorrhagic fever*" OR "Crimean-Congo haemorrhagic fever*" OR Marburg OR MVD OR "Middle East Respiratory syndrome coronavirus" OR "MERS-CoV" OR "MERS Virus*" OR "Middle East respiratory syndrome" OR Merbecovirus*)<br>AND<br>TS=((Africa* NEAR/2 east*) OR Burundi* OR Comoros* OR Djibouti* OR Eritrea* OR Ethiopia* OR Kenya* OR Madagascar* OR Mauritius* OR Rwanda* OR Seychelles* OR Somalia* OR Sudan* OR Tanzania* OR Uganda* OR "Addis Ababa" OR Antananarivo OR Asmara OR Bujumbura OR "Dar es |

| | |
|---|---|
| | Salaam" OR "Dire Dawa" OR "Djibouti City" OR Dodoma OR Gitega OR Hargeisa OR Juba OR Kampala OR Keren OR Khartoum OR Kigali OR Kisumu OR Lindi OR Mahajanga OR Mbarara OR Mogadishu OR Mombasa OR Muhanga OR Moroni OR Muyinga OR Mwanza OR Nairobi OR Ngozi OR Omdurman OR "Port Louis" OR Victoria OR "Great Lakes Region*" OR "Horn of Africa*" OR "Nile Valle*")<br><br>**CAB Direct and CABI Digital Library**<br>("Crimean-congo hemorrhagic fever" OR "Crimean-congo hemorrhagic fevers" OR "Crimean-Congo haemorrhagic fever" OR "Crimean-Congo haemorrhagic fevers" OR CCHF OR "congo virus" OR "congo viruses" OR "Crimean hemorrhagic fever" OR "Crimean hemorrhagic fevers" OR Marburg OR MVD OR "Middle East Respiratory syndrome coronavirus" OR "MERS-CoV" OR "MERS Virus" OR "MERS Viruses" OR "Middle East respiratory syndrome" OR Merbecovirus*)<br>AND<br>("East Africa" OR "Eastern Africa" OR "East African" OR "Eastern African" OR Burundi* OR Comoros* OR Djibouti* OR Eritrea* OR Ethiopia* OR Kenya* OR Madagascar* OR Mauritius* OR Rwanda* OR Seychelles* OR Somalia* OR Sudan* OR Tanzania* OR Uganda* OR "Addis Ababa" OR Antananarivo OR Asmara OR Bujumbura OR "Dar es Salaam" OR "Dire Dawa" OR "Djibouti City" OR Dodoma OR Gitega OR Hargeisa OR Juba OR Kampala OR Keren OR Khartoum OR Kigali OR Kisumu OR Lindi OR Mahajanga OR Mbarara OR Mogadishu OR Mombasa OR Muhanga OR Moroni OR Muyinga OR Mwanza OR Nairobi OR Ngozi OR Omdurman OR "Port Louis" OR Victoria OR "Great Lakes Region" OR "Great Lakes Regions" OR "Horn of Africa" OR "Nile Valle") |
| **2. What risk frameworks and/or methodological tools exist for assessing and managing environmentally driven zoonotic emerging and/or reemerging infectious diseases (EIDs) and what are the gaps in existing biological threat risk analysis frameworks?**<br><br>Inclusion criteria | **Pubmed**<br>**("Zoonoses"[Mesh] OR "Disease Outbreaks"[Mesh:NoExp] OR "Disease Hotspot"[Mesh] OR "Epidemics"[Mesh:NoExp] OR "Space-Time Clustering"[Mesh] OR "Disease Transmission, Infectious"[Mesh] OR "Animals"[Mesh] OR "Livestock"[Mesh] OR "Communicable Diseases, Emerging"[Mesh] OR "Communicable Diseases"[Mesh]** OR Zoono*[tw] OR "animal to human"[tiab:~5] OR "animal to humans"[tiab:~5] OR "animals to human"[tiab:~5] OR "animals to humans"[tiab:~5] OR "infectious disease outbreak"[tiab:~5] OR "infectious disease outbreaks"[tiab:~5] OR "infectious diseases outbreak"[tiab:~5] OR "infectious diseases outbreaks"[tiab:~5] OR epidemic*[tw] OR "disease transmission"[tw] OR "disease spread"[tw] OR "disease cluster*"[tw] OR "cluster of cases"[tw] OR "emerging diseases"[tiab:~5] OR "reemerging diseases"[tiab:~5] OR "re emerging diseases"[tiab:~5] OR spillover*[tw] OR animal*[tw] OR livestock*[tw] OR (viral[tw] AND ("biological threat*"[tw] OR "bio-threat*"[tw])) OR "Hemorrhagic Fever Virus, Crimean-Congo"[Mesh] OR "Crimean-congo hemorrhagic fever*"[tw] OR "Crimean-Congo haemorrhagic fever*"[tw] OR CCHF[tw] OR "congo |

| | |
|---|---|
| - Risk Analysis related<br>- Biological threats<br>- English language | virus*"[tw] OR "Crimean hemorrhagic fever*"[tw] OR "Marburg Virus Disease"[Mesh] OR Marburg[tw] OR MVD[tw] OR "Middle East Respiratory Syndrome Coronavirus"[Mesh] OR "Middle East Respiratory syndrome coronavirus"[tw] OR "MERS-CoV"[tw] OR "MERS Virus*"[tw] OR "Middle East respiratory syndrome"[tw] OR Merbecovirus*[tw] )<br>AND<br>**("Risk Assessment"[Mesh] OR "Decision Making"[Mesh]** OR "risk assessment*"[tw] OR "risk analys*"[tw] OR "risk manag*"[tw] OR "risk communication*"[tw] OR "joint assessment"[tw] OR "risk mitigation"[tw] OR "mitigating risk*"[tw] OR "decision making"[tw] OR "assessment framework*"[tw] OR "analysis framework*"[tw] OR mitigation[tw] OR "integrated risk*"[tw] OR "risks hazard"[tiab:~3] OR "risks hazards"[tiab:~3] OR "risks hazardous"[tiab:~3])<br>AND<br>**("One Health"[Mesh]** OR "one health*"[tw] OR tripartite*[tw] OR "transdisciplinary health*"[tw] OR "trans disciplinary health*"[tw] OR "integrated health*"[tw] OR<br>(("human animal"[tiab:~3] OR "humans animals"[tiab:~3] OR "humans animal"[tiab:~3] OR "human animals"[tiab:~3]) AND (environment*[tw] OR ecosystem*[tw] OR "planetary health"[tw])) OR (("Human agriculture"[tiab:~3] OR "Humans agriculture"[tiab:~3] OR "Human agricultural"[tiab:~3] OR "Humans agricultural"[tiab:~3] OR "Human livestock"[tiab:~3] OR "Humans livestock"[tiab:~3]) AND (environment*[tw] OR ecosystem*[tw] OR "planetary health"[tw])))<br>AND<br>**("Climatic Processes"[Mesh] OR "Ecological and Environmental Phenomena"[Mesh:NoExp] OR "Anthropogenic Effects"[Mesh] OR "Carbon Footprint"[Mesh] OR "Ecosystem"[Mesh] OR "Environment"[Mesh] OR "Environmental Health"[Mesh] OR "Meteorological Concepts"[Mesh] OR "Conservation of Natural Resources"[Mesh]** OR **"Disasters"[Mesh]** OR **"Climate Change"[Mesh] OR "Greenhouse Gases"[Mesh] OR "Ozone Depletion"[Mesh] OR "Carbon Footprint"[Mesh]** OR disaster*[tw] OR postdisaster*[tw] OR posthazard*[tw] OR hazard*[tw] OR avalanche*[tw] OR cyclonic storm*[tw] OR cyclone*[tw] OR hurricane*[tw] OR tropical storm*[tw] OR typhoon*[tw] OR drought*[tw] OR earthquake*[tw] OR flood*[tw] OR landslide*[tw] OR rockslide*[tw] OR mudslide*[tw] OR earth tide*[tw] OR ocean tide*[tw] OR tidalwave*[tw] OR tidal wave*[tw] OR tsunami*[tw] OR tornado*[tw] OR wildfire*[tw] OR wildland fire*[tw] OR brush fire*[tw] OR forest fire*[tw] OR wild fire*[tw] OR facility fire*[tw] OR volcanic*[tw] OR major rainfall*[tw] OR storm[tw] OR storms[tw] OR fire*[tw] OR volcano*[tw] OR volcanic[tw] OR "extreme temperature*"[tw] OR "extreme weather*"[tw] OR heatwave*[tw] OR "heat wave*"[tw] OR "extreme heat*"[tw] OR "extreme cold*"[tw] OR rainfall[tw] OR humidity[tw] OR altitude[tw] OR climate*[tw] OR climatic[tw] OR "global warm*"[tw] OR "greenhouse effect*"[tw] OR "greenhouse gas"[tw] OR "green house effect*"[tw] OR |

| | |
|---|---|
| | "green house gas"[tw] OR "green house gases"[tw] OR "greenhouse gases"[tw] OR "ozone depletion"[tw] OR "ozone hole*"[tw] OR "carbon foot*"[tw] OR deforestation[tw] OR reforestation[tw] OR logging[tw] OR conservation[tw] OR "carrying capacit*"[tw] OR "environmental protection"[tw] OR "environmental biodegradation"[tw] OR "sustainable development*"[tw] OR "smart growth"[tw] OR Meteorological[tw] OR Anthropogenic[tw] OR "human impact*"[tw] OR "land degradation"[tw] OR "forest clearing"[tw] OR "forest restoration"[tw] OR "carbon sink*"[tw] OR "ecosystem respiration"[tw] OR "carbon flux*"[tw] OR environment*[tw] OR livestock*[tw] OR temperature*[tw] OR warm*[tw] OR meteo*[tw] OR ecology[tw] OR ecologically[tw] OR ecological[tw] OR ecosystem**[tw] OR biodiversity[tw] OR "land use"[tw] OR wilderness[tw] OR wildlife[tw] OR forest*[tw])<br><br>**Embase**<br>**('zoonosis'/exp OR 'epidemic'/de OR 'disease hotspot'/exp OR 'spatiotemporal analysis'/exp OR 'zoonotic transmission'/exp OR 'pathogen transmission'/exp OR 'nosocomial transmission'/exp OR 'community transmission'/exp OR 'animal'/exp OR 'livestock'/exp OR 'communicable disease'/exp OR 'emerging infectious disease'/exp OR 'Marburg hemorrhagic fever'/exp OR 'Crimean-Congo hemorrhagic fever virus'/exp OR 'Middle East respiratory syndrome coronavirus'/exp) OR** (Zoono* OR epidemic* OR "disease transmission" OR "disease spread" OR "disease cluster*" OR "cluster of cases" OR (animal* NEAR/5 human*) OR ("infectious disease*" NEAR/5 outbreak*) OR (emerging NEAR/5 disease*) OR (reemerging NEAR/5 disease*) OR spillover* OR animal* OR livestock* OR (viral AND ("biological threat*" OR "bio-threat*")) OR "Crimean-congo hemorrhagic fever*" OR "Crimean-Congo haemorrhagic fever*" OR CCHF OR "congo virus*" OR "Crimean hemorrhagic fever*" OR Marburg OR MVD OR "Middle East Respiratory syndrome coronavirus" OR "MERS-CoV" OR "MERS Virus*" OR "Middle East respiratory syndrome" OR Merbecovirus*):ab,ti,kw<br>AND<br>**('risk assessment'/exp OR 'decision making'/exp)** OR ("risk assessment*" OR "risk analys*" OR "risk manag*" OR "risk communication*" OR "joint assessment" OR "risk mitigation" OR "mitigating risk*" OR "decision making" OR "assessment framework*" OR "analysis framework*" OR mitigation OR "integrated risk*" OR (risks NEAR/3 hazard*)):ab,ti,kw<br>AND<br>**('One Health'/exp)** OR ('one health*':ab,ti,kw OR tripartite*:ab,ti,kw OR 'transdisciplinary health*':ab,ti,kw OR 'trans disciplinary health*':ab,ti,kw OR 'integrated health*':ab,ti,kw) OR (((human* NEAR/3 (animal* OR agricultur* OR livestock*)):ab,ti,kw) AND (environment*:ab,ti,kw OR ecosystem*:ab,ti,kw OR 'planetary health':ab,ti,kw))<br>AND |

| | |
|---|---|
| | **('climate change'/exp OR 'environmental aspects and related phenomena'/de OR 'environmental change'/exp OR 'environmental impact'/exp OR 'human impact (environment)'/exp OR 'carbon footprint'/exp OR 'ecosystem'/exp OR 'environment'/exp OR 'environmental health'/exp OR 'meteorological phenomena'/exp OR 'environmental protection'/exp OR 'disaster'/exp OR 'greenhouse gas emission'/exp OR 'ozone depletion'/exp OR 'carbon footprint'/exp)** OR (disaster* OR postdisaster* OR posthazard* OR hazard* OR avalanche* OR "cyclonic storm*" OR cyclone* OR hurricane* OR "tropical storm*" OR typhoon* OR drought* OR earthquake* OR flood* OR landslide* OR rockslide* OR mudslide* OR "earth tide*" OR "ocean tide*" OR tidalwave* OR "tidal wave*" OR tsunami* OR tornado* OR wildfire* OR "wildland fire*" OR "brush fire*" OR "forest fire*" OR "wild fire*" OR "facility fire*" OR volcanic* OR "major rainfall*" OR storm OR storms OR fire* OR volcano* OR volcanic OR "extreme temperature*" OR "extreme weather*" OR heatwave* OR "heat wave*" OR "extreme heat*" OR "extreme cold*" OR rainfall OR humidity OR altitude OR climate* OR climatic OR "global warm*" OR "greenhouse effect*" OR "greenhouse gas" OR "green house effect*" OR "green house gas" OR "green house gases" OR "greenhouse gases" OR "ozone depletion" OR "ozone hole*" OR "carbon foot*" OR deforestation OR reforestation OR logging OR conservation OR "carrying capacit*" OR "environmental protection" OR "environmental biodegradation" OR "sustainable development*" OR "smart growth" OR Meteorological OR Anthropogenic OR "human impact*" OR "land degradation" OR "forest clearing" OR "forest restoration" OR "carbon sink*" OR "ecosystem respiration" OR "carbon flux*" OR environment* OR livestock* OR temperature* OR warm* OR meteo* OR ecology OR ecologically OR ecological OR ecological OR ecosystem* OR biodiversity OR "land use" OR wilderness OR wildlife OR forest*):ab,ti,kw<br>AND<br>[2000-2023]/py<br><br>**Scopus**<br>TITLE-ABS-KEY((Zoono* OR epidemic* OR "disease transmission" OR "disease spread" OR "disease cluster*" OR "cluster of cases" OR (animal* W/5 human*) OR ("infectious disease*" W/5 outbreak*) OR (emerging W/5 disease*) OR (reemerging W/5 disease*) OR spillover* OR animal* OR livestock* OR (viral AND ("biological threat*" OR "bio-threat*")) OR "Crimean-congo hemorrhagic fever*" OR "Crimean-Congo haemorrhagic fever*" OR CCHF OR "congo virus*" OR "Crimean hemorrhagic fever*" OR Marburg OR MVD OR "Middle East Respiratory syndrome coronavirus" OR "MERS-CoV" OR "MERS Virus*" OR "Middle East respiratory syndrome" OR Merbecovirus*))<br>AND |

| | |
|---|---|
| | TITLE-ABS-KEY ("risk assessment*" OR "risk analys*" OR "risk manag*" OR "risk communication*" OR "joint assessment" OR "risk mitigation" OR "mitigating risk*" OR "decision making" OR "assessment framework*" OR "analysis framework*" OR mitigation OR "integrated risk*" OR (risks W/3 hazard*))<br>AND<br>TITLE-ABS-KEY (("one health*" OR tripartite* OR "transdisciplinary health*" OR "trans disciplinary health*" OR "integrated health*") OR (((human* W/3 (animal* OR agricultur* OR livestock*))) AND (environment* OR ecosystem* OR "planetary health")))<br>AND<br>TITLE-ABS-KEY (disaster* OR postdisaster* OR posthazard* OR hazard* OR avalanche* OR "cyclonic storm*" OR cyclone* OR hurricane* OR "tropical storm*" OR typhoon* OR drought* OR earthquake* OR flood* OR landslide* OR rockslide* OR mudslide* OR "earth tide*" OR "ocean tide*" OR tidalwave* OR "tidal wave*" OR tsunami* OR tornado* OR wildfire* OR "wildland fire*" OR "brush fire*" OR "forest fire*" OR "wild fire*" OR "facility fire*" OR volcanic* OR "major rainfall*" OR storm OR storms OR fire* OR volcano* OR volcanic OR "extreme temperature*" OR "extreme weather*" OR heatwave* OR "heat wave*" OR "extreme heat*" OR "extreme cold*" OR rainfall OR humidity OR altitude OR climate* OR climatic OR "global warm*" OR "greenhouse effect*" OR "greenhouse gas" OR "green house effect*" OR "green house gas" OR "green house gases" OR "greenhouse gases" OR "ozone depletion" OR "ozone hole*" OR "carbon foot*" OR deforestation OR reforestation OR logging OR conservation OR "carrying capacit*" OR "environmental protection" OR "environmental biodegradation" OR "sustainable development*" OR "smart growth" OR Meteorological OR Anthropogenic OR "human impact*" OR "land degradation" OR "forest clearing" OR "forest restoration" OR "carbon sink*" OR "ecosystem respiration" OR "carbon flux*" OR environment* OR livestock* OR temperature* OR warm* OR meteo* OR ecology OR ecologically OR ecological OR ecosystem* OR biodiversity OR "land use" OR wilderness OR wildlife OR forest*)<br>AND<br>( PUBYEAR > 1999 ) AND NOT ( INDEX ( medline ) OR INDEX ( embase ) )<br><br>**Web of Science**<br>TS= ((Zoono* OR epidemic* OR "disease transmission" OR "disease spread" OR "disease cluster*" OR "cluster of cases" OR (animal* NEAR/5 human*) OR ("infectious disease*" NEAR/5 outbreak*) OR (emerging NEAR/5 disease*) OR (reemerging NEAR/5 disease*) OR spillover* OR animal* OR livestock* OR (viral AND ("biological threat*" OR "bio-threat*")) OR "Crimean-congo hemorrhagic fever*" OR "Crimean-Congo haemorrhagic fever*" OR CCHF OR "congo virus*" OR "Crimean |

| | |
|---|---|
| | hemorrhagic fever*" OR Marburg OR MVD OR "Middle East Respiratory syndrome coronavirus" OR "MERS-CoV" OR "MERS Virus*" OR "Middle East respiratory syndrome" OR Merbecovirus*))<br>AND<br>TS= ("risk assessment*" OR "risk analys*" OR "risk manag*" OR "risk communication*" OR "joint assessment" OR "risk mitigation" OR "mitigating risk*" OR "decision making" OR "assessment framework*" OR "analysis framework*" OR mitigation OR "integrated risk*" OR (risks NEAR/3 hazard*))<br>AND<br>TS= (("one health*" OR tripartite* OR "transdisciplinary health*" OR "trans disciplinary health*" OR "integrated health*") OR (((human* NEAR/3 (animal* OR agricultur* OR livestock*))) AND (environment* OR ecosystem* OR "planetary health")))<br>AND<br>TS= (disaster* OR postdisaster* OR posthazard* OR hazard* OR avalanche* OR "cyclonic storm*" OR cyclone* OR hurricane* OR "tropical storm*" OR typhoon* OR drought* OR earthquake* OR flood* OR landslide* OR rockslide* OR mudslide* OR "earth tide*" OR "ocean tide*" OR tidalwave* OR "tidal wave*" OR tsunami* OR tornado* OR wildfire* OR "wildland fire*" OR "brush fire*" OR "forest fire*" OR "wild fire*" OR "facility fire*" OR volcanic* OR "major rainfall*" OR storm OR storms OR fire* OR volcano* OR volcanic OR "extreme temperature*" OR "extreme weather*" OR heatwave* OR "heat wave*" OR "extreme heat*" OR "extreme cold*" OR rainfall OR humidity OR altitude OR climate* OR climatic OR "global warm*" OR "greenhouse effect*" OR "greenhouse gas" OR "green house effect*" OR "green house gas" OR "green house gases" OR "greenhouse gases" OR "ozone depletion" OR "ozone hole*" OR "carbon foot*" OR deforestation OR reforestation OR logging OR conservation OR "carrying capacit*" OR "environmental protection" OR "environmental biodegradation" OR "sustainable development*" OR "smart growth" OR Meteorological OR Anthropogenic OR "human impact*" OR "land degradation" OR "forest clearing" OR "forest restoration" OR "carbon sink*" OR "ecosystem respiration" OR "carbon flux*" OR environment* OR livestock* OR temperature* OR warm* OR meteo* OR ecology OR ecologically OR ecological OR ecosystem* OR biodiversity OR "land use" OR wilderness OR wildlife OR forest*)<br><br>**CABI Digital Library**<br>Title:(Zoono* OR epidemic* OR "disease transmission" OR "disease spread" OR "disease cluster" OR "disease clusters" OR "cluster of cases" OR "animal human" OR "animals human" OR "animal humans" OR "animals humans" OR "infectious disease outbreak" OR "infectious disease outbreaks" OR "infectious diseases outbreak" OR "infectious diseases outbreaks" OR "emerging disease" OR "emerging diseases" OR |

| | |
|---|---|
| | "reemerging disease" OR "reemerging diseases" OR spillover* OR animal* OR livestock* OR "Crimean-congo hemorrhagic fever" OR "Crimean-congo hemorrhagic fevers" OR "Crimean-Congo haemorrhagic fever" OR "Crimean-Congo haemorrhagic fevers" OR CCHF OR "congo virus" OR "congo viruses" OR "Crimean hemorrhagic fever" OR "Crimean hemorrhagic fevers" OR Marburg OR MVD OR "Middle East Respiratory syndrome coronavirus" OR "MERS-CoV" OR "MERS Virus" OR "MERS Viruses" OR "Middle East respiratory syndrome" OR Merbecovirus*) OR Title:(viral AND ("biological threat" OR "biological threats" OR "bio-threat" OR "bio-threats")) OR ab:(Zoono* OR epidemic* OR "disease transmission" OR "disease spread" OR "disease cluster" OR "disease clusters" OR "cluster of cases" OR "animal human" OR "animals human" OR "animal humans" OR "animals humans" OR "infectious disease outbreak" OR "infectious disease outbreaks" OR "infectious diseases outbreak" OR "infectious diseases outbreaks" OR "emerging disease" OR "emerging diseases" OR "reemerging disease" OR "reemerging diseases" OR spillover* OR animal* OR livestock* OR "Crimean-congo hemorrhagic fever" OR "Crimean-congo hemorrhagic fevers" OR "Crimean-Congo haemorrhagic fever" OR "Crimean-Congo haemorrhagic fevers" OR CCHF OR "congo virus" OR "congo viruses" OR "Crimean hemorrhagic fever" OR "Crimean hemorrhagic fevers" OR Marburg OR MVD OR "Middle East Respiratory syndrome coronavirus" OR "MERS-CoV" OR "MERS Virus" OR "MERS Viruses" OR "Middle East respiratory syndrome" OR Merbecovirus*) OR ab:(viral AND ("biological threat" OR "biological threats" OR "bio-threat" OR "bio-threats")) OR indexingterm:(Zoono* OR epidemic* OR "disease transmission" OR "disease spread" OR "disease cluster" OR "disease clusters" OR "cluster of cases" OR "animal human" OR "animals human" OR "animal humans" OR "animals humans" OR "infectious disease outbreak" OR "infectious disease outbreaks" OR "infectious diseases outbreak" OR "infectious diseases outbreaks" OR "emerging disease" OR "emerging diseases" OR "reemerging disease" OR "reemerging diseases" OR spillover* OR animal* OR livestock* OR "Crimean-congo hemorrhagic fever" OR "Crimean-congo hemorrhagic fevers" OR "Crimean-Congo haemorrhagic fever" OR "Crimean-Congo haemorrhagic fevers" OR CCHF OR "congo virus" OR "congo viruses" OR "Crimean hemorrhagic fever" OR "Crimean hemorrhagic fevers" OR Marburg OR MVD OR "Middle East Respiratory syndrome coronavirus" OR "MERS-CoV" OR "MERS Virus" OR "MERS Viruses" OR "Middle East respiratory syndrome" OR Merbecovirus*) OR indexing:(viral AND ("biological threat" OR "biological threats" OR "bio-threat" OR "bio-threats"))<br>AND<br>Title:("risk assessment" OR "risk assessments" OR "risk analysis" OR "risk analyses" OR "risk management" OR "risk communication" OR "joint assessment" OR "risk mitigation" OR "mitigating risk" OR "mitigating risks" OR "decision making" OR "assessment framework" OR "assessment frameworks" OR "analysis framework" OR "analysis frameworks" OR mitigation OR "integrated risk" OR "integrated |

| | |
|---|---|
| | risks" OR "risk hazard" OR "risk hazards" OR "risks hazard" OR "risks hazards") OR ab:("risk assessment" OR "risk assessments" OR "risk analysis" OR "risk analyses" OR "risk management" OR "risk communication" OR "joint assessment" OR "risk mitigation" OR "mitigating risk" OR "mitigating risks" OR "decision making" OR "assessment framework" OR "assessment frameworks" OR "analysis framework" OR "analysis frameworks" OR mitigation OR "integrated risk" OR "integrated risks" OR "risk hazard" OR "risk hazards" OR "risks hazard" OR "risks hazards") OR indexingterm:("risk assessment" OR "risk assessments" OR "risk analysis" OR "risk analyses" OR "risk management" OR "risk communication" OR "joint assessment" OR "risk mitigation" OR "mitigating risk" OR "mitigating risks" OR "decision making" OR "assessment framework" OR "assessment frameworks" OR "analysis framework" OR "analysis frameworks" OR mitigation OR "integrated risk" OR "integrated risks" OR "risk hazard" OR "risk hazards" OR "risks hazard" OR "risks hazards")<br>AND<br>Title:("one health" OR "one healthy" OR tripartite* OR "transdisciplinary health" OR "transdisciplinary healthy" OR "trans disciplinary health" OR "trans disciplinary healthy" OR "integrated health" OR "integrated healthy") OR title:(((human* AND (animal* OR agricultur* OR livestock*))) AND (environment* OR ecosystem* OR "planetary health")) OR ab:("one health" OR "one healthy" OR tripartite* OR "transdisciplinary health" OR "transdisciplinary healthy" OR "trans disciplinary health" OR "trans disciplinary healthy" OR "integrated health" OR "integrated healthy") OR ab:(((human* AND (animal* OR agricultur* OR livestock*))) AND (environment* OR ecosystem* OR "planetary health")) OR indexingterm:("one health" OR "one healthy" OR tripartite* OR "transdisciplinary health" OR "transdisciplinary healthy" OR "trans disciplinary health" OR "trans disciplinary healthy" OR "integrated health" OR "integrated healthy") OR indexingterm:(((human* AND (animal* OR agricultur* OR livestock*))) AND (environment* OR ecosystem* OR "planetary health"))<br>AND<br>Title:(disaster* OR postdisaster* OR posthazard* OR hazard* OR avalanche* OR cyclone* OR hurricane* OR typhoon* OR drought* OR earthquake* OR flood* OR landslide* OR rockslide* OR mudslide* OR "earth tide" OR "earth tides" OR "ocean tide" OR "ocean tides" OR tidalwave* OR "tidal wave" OR "tidal waves" OR tsunami* OR tornado* OR wildfire* OR volcanic* OR "major rainfall" OR "major rainfalls" OR storm OR storms OR fire* OR volcano* OR "extreme temperature" OR "extreme temperatures" OR "extreme weather" OR "extreme weathers" OR heatwave* OR "heat wave" OR "heat waves" OR "extreme heat" OR "extreme heats" OR "extreme cold" OR rainfall OR humidity OR altitude OR climate* OR climatic OR "global warm" OR "global warming" OR "greenhouse effect" OR "greenhouse effects" OR "greenhouse gas" OR "green house effect" OR "green house effects" OR "green house gas" OR "green house gases" OR "greenhouse gases" OR "ozone depletion" OR "ozone hole" OR "ozone holes" OR |

| | |
|---|---|
| | "carbon foot" OR "carbon footprint" OR "carbon footprints" OR deforestation OR reforestation OR logging OR conservation OR "carrying capacity" OR "carrying capacities" OR "environmental protection" OR "environmental biodegradation" OR "sustainable development" OR "sustainable developments" OR "smart growth" OR Meteorological OR Anthropogenic OR "human impact" OR "human impacts" OR "land degradation" OR "forest clearing" OR "forest restoration" OR "carbon sink" OR "carbon sinks" OR "carbon sinking" OR "ecosystem respiration" OR "carbon flux" OR environment* OR livestock* OR temperature* OR warm* OR meteo* OR ecology OR ecologically OR ecological OR ecosystem* OR biodiversity OR "land use" OR wilderness OR wildlife OR forest*) OR ab:(disaster* OR postdisaster* OR posthazard* OR hazard* OR avalanche* OR cyclone* OR hurricane* OR typhoon* OR drought* OR earthquake* OR flood* OR landslide* OR rockslide* OR mudslide* OR "earth tide" OR "earth tides" OR "ocean tide" OR "ocean tides" OR tidalwave* OR "tidal wave" OR "tidal waves" OR tsunami* OR tornado* OR wildfire* OR volcanic* OR "major rainfall" OR "major rainfalls" OR storm OR storms OR fire* OR volcano* OR "extreme temperature" OR "extreme temperatures" OR "extreme weather" OR "extreme weathers" OR heatwave* OR "heat wave" OR "heat waves" OR "extreme heat" OR "extreme heats" OR "extreme cold" OR rainfall OR humidity OR altitude OR climate* OR climatic OR "global warm" OR "global warming" OR "greenhouse effect" OR "greenhouse effects" OR "greenhouse gas" OR "green house effect" OR "green house effects" OR "green house gas" OR "green house gases" OR "greenhouse gases" OR "ozone depletion" OR "ozone hole" OR "ozone holes" OR "carbon foot" OR "carbon footprint" OR "carbon footprints" OR deforestation OR reforestation OR logging OR conservation OR "carrying capacity" OR "carrying capacities" OR "environmental protection" OR "environmental biodegradation" OR "sustainable development" OR "sustainable developments" OR "smart growth" OR Meteorological OR Anthropogenic OR "human impact" OR "human impacts" OR "land degradation" OR "forest clearing" OR "forest restoration" OR "carbon sink" OR "carbon sinks" OR "carbon sinking" OR "ecosystem respiration" OR "carbon flux" OR environment* OR livestock* OR temperature* OR warm* OR meteo* OR ecology OR ecologically OR ecological OR ecosystem* OR biodiversity OR "land use" OR wilderness OR wildlife OR forest*) OR indexingterm:(disaster* OR postdisaster* OR posthazard* OR hazard* OR avalanche* OR cyclone* OR hurricane* OR typhoon* OR drought* OR earthquake* OR flood* OR landslide* OR rockslide* OR mudslide* OR "earth tide" OR "earth tides" OR "ocean tide" OR "ocean tides" OR tidalwave* OR "tidal wave" OR "tidal waves" OR tsunami* OR tornado* OR wildfire* OR volcanic* OR "major rainfall" OR "major rainfalls" OR storm OR storms OR fire* OR volcano* OR "extreme temperature" OR "extreme temperatures" OR "extreme weather" OR "extreme weathers" OR heatwave* OR "heat wave" OR "heat waves" OR "extreme heat" OR "extreme heats" OR "extreme cold" OR rainfall OR humidity OR altitude OR climate* OR climatic OR "global warm" OR "global warming" OR "greenhouse effect" OR "greenhouse effects" OR "greenhouse gas" OR "green house effect" OR "green |

| | |
|---|---|
| | house effects" OR "green house gas" OR "green house gases" OR "greenhouse gases" OR "ozone depletion" OR "ozone hole" OR "ozone holes" OR "carbon foot" OR "carbon footprint" OR "carbon footprints" OR deforestation OR reforestation OR logging OR conservation OR "carrying capacity" OR "carrying capacities" OR "environmental protection" OR "environmental biodegradation" OR "sustainable development" OR "sustainable developments" OR "smart growth" OR Meteorological OR Anthropogenic OR "human impact" OR "human impacts" OR "land degradation" OR "forest clearing" OR "forest restoration" OR "carbon sink" OR "carbon sinks" OR "carbon sinking" OR "ecosystem respiration" OR "carbon flux" OR environment* OR livestock* OR temperature* OR warm* OR meteo* OR ecology OR ecologically OR ecological OR ecosystem* OR biodiversity OR "land use" OR wilderness OR wildlife OR forest*)<br><br>CAB Direct<br>(Zoono* OR epidemic* OR "disease transmission" OR "disease spread" OR "disease cluster*" OR "cluster of cases" OR "animal* human*" OR "infectious disease* outbreak*" OR "emerging disease*" OR "reemerging disease*" OR spillover* OR animal* OR livestock* OR (viral AND ("biological threat*" OR "bio-threat*")) OR "Crimean-congo hemorrhagic fever*" OR "Crimean-Congo haemorrhagic fever*" OR CCHF OR "congo virus*" OR "Crimean hemorrhagic fever*" OR Marburg OR MVD OR "Middle East Respiratory syndrome coronavirus" OR "MERS-CoV" OR "MERS Virus*" OR "Middle East respiratory syndrome" OR Merbecovirus*)<br>AND<br>("risk assessment*" OR "risk analys*" OR "risk manag*" OR "risk communication*" OR "joint assessment" OR "risk mitigation" OR "mitigating risk*" OR "decision making" OR "assessment framework*" OR "analysis framework*" OR mitigation OR "integrated risk*" OR "risk* hazard*")<br>AND<br>("one health*" OR tripartite* OR "transdisciplinary health*" OR "trans disciplinary health*" OR "integrated health*") OR (((human* AND (animal* OR agricultur* OR livestock*))) AND (environment* OR ecosystem* OR "planetary health"))<br>AND<br>(disaster* OR postdisaster* OR posthazard* OR hazard* OR avalanche* OR cyclone* OR hurricane* OR typhoon* OR drought* OR earthquake* OR flood* OR landslide* OR rockslide* OR mudslide* OR "earth tide*" OR "ocean tide*" OR tidalwave* OR "tidal wave*" OR tsunami* OR tornado* OR wildfire* OR volcanic* OR "major rainfall*" OR storm OR storms OR fire* OR volcano* OR "extreme temperature*" OR "extreme weather*" OR heatwave* OR "heat wave*" OR "extreme heat*" OR "extreme cold*" OR rainfall OR humidity OR altitude OR climate* OR climatic OR "global warm*" OR "greenhouse effect*" |

| | |
|---|---|
| | OR "greenhouse gas" OR "green house effect*" OR "green house gas" OR "green house gases" OR "greenhouse gases" OR "ozone depletion" OR "ozone hole*" OR "carbon foot*" OR deforestation OR reforestation OR logging OR conservation OR "carrying capacit*" OR "environmental protection" OR "environmental biodegradation" OR "sustainable development*" OR "smart growth" OR Meteorological OR Anthropogenic OR "human impact*" OR "land degradation" OR "forest clearing" OR "forest restoration" OR "carbon sink*" OR "ecosystem respiration" OR "carbon flux*" OR environment* OR livestock* OR temperature* OR warm* OR meteo* OR ecology OR ecologically OR ecological OR ecosystem* OR biodiversity OR "land use" OR wilderness OR wildlife OR forest*) |
| **3. What types of policy recommendations applicable to the East Africa region are in place to address environmental drivers of emerging/reemerging zoonotic biological threats such as CCHFV, MARV, and MERS-CoV?**<br><br>Inclusion criteria<br>- Policy recommendations<br>- Recommendations could apply to East Africa<br>- English Language | **Pubmed**<br>**("Zoonoses"[Mesh] OR "Disease Outbreaks"[Mesh:NoExp] OR "Disease Hotspot"[Mesh] OR "Epidemics"[Mesh:NoExp] OR "Space-Time Clustering"[Mesh] OR "Disease Transmission, Infectious"[Mesh]** OR **"Animals"[Mesh] OR "Livestock"[Mesh] OR "Communicable Diseases, Emerging"[Mesh] OR "Communicable Diseases"[Mesh]** OR Zoono*[tw] OR "animal to human"[tiab:~5] OR "animal to humans"[tiab:~5] OR "animals to human"[tiab:~5] OR "animals to humans"[tiab:~5] OR "infectious disease outbreak"[tiab:~5] OR "infectious disease outbreaks"[tiab:~5] OR "infectious diseases outbreak"[tiab:~5] OR "infectious diseases outbreaks"[tiab:~5] OR epidemic*[tw] OR "disease transmission"[tw] OR "disease spread"[tw] OR "disease cluster*"[tw] OR "cluster of cases"[tw] OR "emerging diseases"[tiab:~5] OR "reemerging diseases"[tiab:~5] OR "re emerging diseases"[tiab:~5] OR spillover*[tw] OR animal*[tw] OR livestock*[tw] OR (viral[tw] AND ("biological threat*"[tw] OR "bio-threat*"[tw])) OR **"Hemorrhagic Fever Virus, Crimean-Congo"[Mesh]** OR "Crimean-congo hemorrhagic fever*"[tw] OR CCHF[tw] OR "congo virus*"[tw] OR "Crimean hemorrhagic fever*"[tw] OR **"Marburg Virus Disease"[Mesh]** OR Marburg[tw] OR MVD[tw] OR **"Middle East Respiratory Syndrome Coronavirus"[Mesh]** OR "Middle East Respiratory syndrome coronavirus"[tw] OR "MERS-CoV"[tw] OR "MERS Virus*"[tw] OR "Middle East respiratory syndrome"[tw] OR Merbecovirus*[tw])<br>**("Africa, Eastern"[Mesh]** OR "Africa east"[tiab:~2] OR "Africa eastern"[tiab:~2] OR "African east"[tiab:~2] OR "African eastern"[tiab:~2] OR "africans east"[tiab:~2] OR "africans eastern"[tiab:~2] OR Burundi*[tw] OR Comoros*[tw] OR Djibouti*[tw] OR Eritrea*[tw] OR Ethiopia*[tw] OR Kenya*[tw] OR Madagascar*[tw] OR Mauritius* [tw] OR Rwanda*[tw] OR Seychelles*[tw] OR Somalia*[tw] OR Sudan*[tw] OR Tanzania*[tw] OR Uganda*[tw] OR "Addis Ababa"[tw] OR Antananarivo[tw] OR Asmara[tw] OR Bujumbura[tw] OR "Dar es Salaam"[tw] OR "Dire Dawa"[tw] OR "Djibouti City"[tw] OR Dodoma[tw] OR Gitega[tw] OR Hargeisa[tw] OR Juba[tw] OR Kampala[tw] OR Keren[tw] OR Khartoum[tw] OR Kigali[tw] OR Kisumu[tw] OR Lindi[tw] OR Mahajanga[tw] OR Mbarara[tw] OR Mogadishu[tw] OR Mombasa[tw] OR Muhanga[tw] OR Moroni[tw] OR Muyinga[tw] OR Mwanza[tw] |

| | |
|---|---|
| | OR Nairobi[tw] OR Ngozi[tw] OR Omdurman[tw] OR "Port Louis"[tw] OR Victoria[tw] OR "Great Lakes Region*"[tw] OR "Horn of Africa*"[tw] OR "Nile Valle*"[tw])<br>AND<br>**("Climate"[Mesh] OR "Ecosystem"[Mesh] OR "Natural Resources"[Mesh] OR "Disasters"[Mesh:NoExp] OR "Disaster Planning"[Mesh:NoExp] OR "Natural Disasters"[Mesh] OR "Environmental Health"[Mesh:NoExp] OR "Sanitation"[Mesh:NoExp] OR "Conservation of Natural Resources"[Mesh] OR "Wastewater"[Mesh] OR "Waste Management"[Mesh:NoExp] OR** ecosystem*[tw] OR ecological*[tw] OR climate*[tw] OR "natural resources"[tw] OR "disaster plan*"[tw] OR "natural disaster*"[tw] OR "environmental health"[tw] OR "environmental protection"[tw] **) OR** ((**"Environment"[Mesh] OR** environment*[tw]) AND (drivers[tw] OR factors[tw]))<br>AND<br>(**"Policy"[Mesh:NoExp] OR "Public Policy"[Mesh:NoExp] OR "Environmental Policy"[Mesh:NoExp] OR "Health Policy"[Mesh:NoExp] OR "International Health Regulations"[Mesh] OR "Sustainable Development"[Mesh] OR "Policy Making"[Mesh:NoExp] OR "Government Regulation"[Mesh] OR "Legislation as Topic"[Mesh:NoExp] OR "Decision Making, Shared"[Mesh] OR "Stakeholder Participation"[Mesh] OR "Health Planning Guidelines"[Mesh] OR "Health Priorities"[Mesh] OR "Health Planning"[Mesh:NoExp]** OR Policy change*[tw] OR Policy problem*[tw] OR Policy forecast*[tw] OR Policy solution*[tw] OR Policy stud*[tw] OR Policy environment*[tw] OR Policy development*[tw] OR Policy option*[tw] OR Policy alternative*[tw] OR Policy recommendation*[tw] OR policy implement*[tw] OR Policy maker*[tw] OR Decision maker*[tw] OR "National government*"[tw] OR Policy adoption[tw] OR Policy evaluation*[tw] OR Policy outcome*[tw] OR Policy performance[tw] OR "Health policy and planning"[tw] OR "policy analysis"[tw] OR "Policy formulation*"[tw] OR Policy brief*[tw] OR Policy reform*[tw] OR Public policy[tw] OR public policies[tw] OR Stakeholder engagement*[tw] OR Health polic*[tw] OR "Evidence-based policy"[tw] OR Evidence-based policies*[tw] OR "Evidence-informed policy"[tw] OR "Evidence-informed policies"[tw] OR "cross sectoral decision making"[tiab:~5] OR "risk informed decision making"[tw] OR "government authorit*"[tw] OR "health planning"[tw] OR "Health care reform*"[tw] OR Regulation*[tw] OR "Land management"[tw] OR "Land policy"[tw] OR "land policies"[tw] OR "Policy engagement"[tw] OR "Policy environment*"[tw])<br><br>**Embase**<br>**('zoonosis'/exp OR 'epidemic'/de OR 'disease hotspot'/exp OR 'spatiotemporal analysis'/exp OR 'zoonotic transmission'/exp OR 'pathogen transmission'/exp OR 'nosocomial transmission'/exp OR 'community transmission'/exp OR 'animal'/exp OR 'livestock'/exp OR 'communicable disease'/exp** |

| | |
|---|---|
| | **OR 'emerging infectious disease'/exp OR 'Marburg hemorrhagic fever'/exp OR 'Crimean-Congo hemorrhagic fever virus'/exp OR 'Middle East respiratory syndrome coronavirus'/exp) OR** (Zoono* OR epidemic* OR "disease transmission" OR "disease spread" OR "disease cluster*" OR "cluster of cases" OR (animal* NEAR/5 human*) OR ("infectious disease*" NEAR/5 outbreak*) OR (emerging NEAR/5 disease*) OR (reemerging NEAR/5 disease*) OR spillover* OR animal* OR livestock* OR (viral AND ("biological threat*" OR "bio-threat*")) OR "Crimean-congo hemorrhagic fever*" OR CCHF OR "congo virus*" OR "Crimean hemorrhagic fever*" OR Marburg OR MVD OR "Middle East Respiratory syndrome coronavirus" OR "MERS-CoV" OR "MERS Virus*" OR "Middle East respiratory syndrome" OR Merbecovirus*):ab,ti,kw<br>AND<br>**'east african'/exp** OR ((Africa* NEAR/2 east*) OR Burundi* OR Comoros* OR Djibouti* OR Eritrea* OR Ethiopia* OR Kenya* OR Madagascar* OR Mauritius* OR Rwanda* OR Seychelles* OR Somalia* OR Sudan* OR Tanzania* OR Uganda* OR "Addis Ababa" OR Antananarivo OR Asmara OR Bujumbura OR "Dar es Salaam" OR "Dire Dawa" OR "Djibouti City" OR Dodoma OR Gitega OR Hargeisa OR Juba OR Kampala OR Keren OR Khartoum OR Kigali OR Kisumu OR Lindi OR Mahajanga OR Mbarara OR Mogadishu OR Mombasa OR Muhanga OR Moroni OR Muyinga OR Mwanza OR Nairobi OR Ngozi OR Omdurman OR "Port Louis" OR Victoria OR "Great Lakes Region*" OR "Horn of Africa*" OR "Nile Valle*"):ab,ti,kw<br>AND<br>**('climate'/exp OR 'ecosystem'/exp OR 'natural resource'/exp OR 'disaster'/de OR 'natural disaster'/exp OR 'disaster planning'/de OR 'disaster mitigation'/exp OR 'disaster preparedness'/exp OR 'environmental health'/de OR 'sanitation'/de OR 'pollution control'/exp OR 'environmental protection'/exp OR 'waste management'/de OR 'sewage treatment'/exp)** OR ("environmental health" OR "environmental protection" OR ecosystem* OR ecological* OR climate* OR "natural resources" OR "disaster plan*" OR "natural disaster*"):ab,ti,kw OR ((**'environment'/exp**) AND (drivers OR factors):ab,ti,kw) OR ((environment*:ab,ti,kw) AND (drivers OR factors)):ab,ti,kw<br>AND<br>**('policy'/de OR 'public policy'/de OR 'environmental policy'/de OR 'health care policy'/de OR 'international health regulation'/exp OR 'sustainable development'/exp OR 'management'/de OR 'government regulation'/exp OR 'law'/de OR 'shared decision making'/exp OR 'stakeholder engagement'/exp OR 'health care planning'/exp)** OR ("Policy change*" OR "Policy problem*" OR "Policy forecast*" OR "Policy solution*" OR "Policy stud*" OR "Policy environment*" OR "Policy development*" OR "Policy option*" OR "Policy alternative*" OR "Policy recommendation*" OR "policy implement*" OR "Policy maker*" OR "Decision maker*" OR "National government*" OR "Policy |

| | |
|---|---|
| | adoption" OR "Policy evaluation*" OR "Policy outcome*" OR "Policy performance" OR "Health policy and planning" OR "policy analysis" OR "Policy formulation*" OR "Policy brief*" OR "Policy reform*" OR "Public policy" OR "public policies" OR "Stakeholder engagement*" OR "Health polic*" OR "Evidence-based policy" OR "Evidence-based policies*" OR "Evidence-informed policy" OR "Evidence-informed policies" OR ("cross sectoral" NEAR/5 "decision making") OR "risk informed decision making" OR "government authorit*" OR "health planning" OR "Health care reform*" OR Regulation* OR "Land management" OR "Land policy" OR "land policies" OR "Policy engagement" OR "Policy environment*"):ab,ti,kw<br>AND<br>[2000-2023]/py<br><br>**Scopus**<br>TITLE-ABS-KEY((Zoono* OR epidemic* OR "disease transmission" OR "disease spread" OR "disease cluster*" OR "cluster of cases" OR (animal* W/5 human*) OR ("infectious disease*" W/5 outbreak*) OR (emerging W/5 disease*) OR (reemerging W/5 disease*) OR spillover* OR animal* OR livestock* OR (viral AND ("biological threat*" OR "bio-threat*")) OR "Crimean-congo hemorrhagic fever*" OR CCHF OR "congo virus*" OR "Crimean hemorrhagic fever*" OR Marburg OR MVD OR "Middle East Respiratory syndrome coronavirus" OR "MERS-CoV" OR "MERS Virus*" OR "Middle East respiratory syndrome" OR Merbecovirus*))<br>AND<br>TITLE-ABS-KEY ((Africa* W/2 east*) OR Burundi* OR Comoros* OR Djibouti* OR Eritrea* OR Ethiopia* OR Kenya* OR Madagascar* OR Mauritius* OR Rwanda* OR Seychelles* OR Somalia* OR Sudan* OR Tanzania* OR Uganda* OR "Addis Ababa" OR Antananarivo OR Asmara OR Bujumbura OR "Dar es Salaam" OR "Dire Dawa" OR "Djibouti City" OR Dodoma OR Gitega OR Hargeisa OR Juba OR Kampala OR Keren OR Khartoum OR Kigali OR Kisumu OR Lindi OR Mahajanga OR Mbarara OR Mogadishu OR Mombasa OR Muhanga OR Moroni OR Muyinga OR Mwanza OR Nairobi OR Ngozi OR Omdurman OR "Port Louis" OR Victoria OR "Great Lakes Region*" OR "Horn of Africa*" OR "Nile Valle*")<br>AND<br>TITLE-ABS-KEY (ecosystem* OR ecological* OR climate* OR "natural resources" OR "disaster plan*" OR "natural disaster*" OR "environmental health" OR "environmental protection") OR TITLE-ABS-KEY(environment* AND (drivers OR factors))<br>AND |

| | |
|---|---|
| | TITLE-ABS-KEY ("Policy change*" OR "Policy problem*" OR "Policy forecast*" OR "Policy solution*" OR "Policy stud*" OR "Policy environment*" OR "Policy development*" OR "Policy option*" OR "Policy alternative*" OR "Policy recommendation*" OR "policy implement*" OR "Policy maker*" OR "Decision maker*" OR "National government*" OR "Policy adoption" OR "Policy evaluation*" OR "Policy outcome*" OR "Policy performance" OR "Health policy and planning" OR "policy analysis" OR "Policy formulation*" OR "Policy brief*" OR "Policy reform*" OR "Public policy" OR "public policies" OR "Stakeholder engagement*" OR "Health polic*" OR "Evidence-based policy" OR "Evidence-based policies*" OR "Evidence-informed policy" OR "Evidence-informed policies" OR ("cross sectoral" W/5 "decision making") OR "risk informed decision making" OR "government authorit*" OR "health planning" OR "Health care reform*" OR Regulation* OR "Land management" OR "Land policy" OR "land policies" OR "Policy engagement" OR "Policy environment*")<br>AND<br>( PUBYEAR > 1999 ) AND NOT ( INDEX ( medline ) OR INDEX ( embase ) )<br><br>**Web of Science**<br>TS= ((Zoono* OR epidemic* OR "disease transmission" OR "disease spread" OR "disease cluster*" OR "cluster of cases" OR (animal* NEAR/5 human*) OR ("infectious disease*" NEAR/5 outbreak*) OR (emerging NEAR/5 disease*) OR (reemerging NEAR/5 disease*) OR spillover* OR animal* OR livestock* OR (viral AND ("biological threat*" OR "bio-threat*")) OR "Crimean-congo hemorrhagic fever*" OR CCHF OR "congo virus*" OR "Crimean hemorrhagic fever*" OR Marburg OR MVD OR "Middle East Respiratory syndrome coronavirus" OR "MERS-CoV" OR "MERS Virus*" OR "Middle East respiratory syndrome" OR Merbecovirus*))<br>AND<br>TS= ((Zoono* OR epidemic* OR "disease transmission" OR "disease spread" OR "disease cluster*" OR "cluster of cases" OR (animal* NEAR/5 human*) OR ("infectious disease*" NEAR/5 outbreak*) OR (emerging NEAR/5 disease*) OR (reemerging NEAR/5 disease*) OR spillover* OR animal* OR livestock* OR (viral AND ("biological threat*" OR "bio-threat*")) OR "Crimean-congo hemorrhagic fever*" OR CCHF OR "congo virus*" OR "Crimean hemorrhagic fever*" OR Marburg OR MVD OR "Middle East Respiratory syndrome coronavirus" OR "MERS-CoV" OR "MERS Virus*" OR "Middle East respiratory syndrome" OR Merbecovirus*))<br>AND<br>TS=((Africa* NEAR/2 east*) OR Burundi* OR Comoros* OR Djibouti* OR Eritrea* OR Ethiopia* OR Kenya* OR Madagascar* OR Mauritius* OR Rwanda* OR Seychelles* OR Somalia* OR Sudan* OR Tanzania* OR Uganda* OR "Addis Ababa" OR Antananarivo OR Asmara OR Bujumbura OR "Dar es |

| | |
|---|---|
| | Salaam" OR "Dire Dawa" OR "Djibouti City" OR Dodoma OR Gitega OR Hargeisa OR Juba OR Kampala OR Keren OR Khartoum OR Kigali OR Kisumu OR Lindi OR Mahajanga OR Mbarara OR Mogadishu OR Mombasa OR Muhanga OR Moroni OR Muyinga OR Mwanza OR Nairobi OR Ngozi OR Omdurman OR "Port Louis" OR Victoria OR "Great Lakes Region*" OR "Horn of Africa*" OR "Nile Valle*")<br>AND<br>TS= (environment* OR ecosystem* OR ecological* OR climate* OR "natural resources" OR "disaster plan*" OR "natural disaster*" OR "environmental health" OR "environmental protection") OR TS=(environment* AND (drivers OR factors))<br>AND<br>TS= ("Policy change*" OR "Policy problem*" OR "Policy forecast*" OR "Policy solution*" OR "Policy stud*" OR "Policy environment*" OR "Policy development*" OR "Policy option*" OR "Policy alternative*" OR "Policy recommendation*" OR "policy implement*" OR "Policy maker*" OR "Decision maker*" OR "National government*" OR "Policy adoption" OR "Policy evaluation*" OR "Policy outcome*" OR "Policy performance" OR "Health policy and planning" OR "policy analysis" OR "Policy formulation*" OR "Policy brief*" OR "Policy reform*" OR "Public policy" OR "public policies" OR "Stakeholder engagement*" OR "Health polic*" OR "Evidence-based policy" OR "Evidence-based policies*" OR "Evidence-informed policy" OR "Evidence-informed policies" OR ("cross sectoral" NEAR/5 "decision making") OR "risk informed decision making" OR "government authorit*" OR "health planning" OR "Health care reform*" OR Regulation* OR "Land management" OR "Land policy" OR "land policies" OR "Policy engagement" OR "Policy environment*")<br>AND<br>PY=(2000 OR 2001 OR 2002 OR 2003 OR 2004 OR 2005 OR 2006 OR 2007 OR 2008 OR 2009 OR 2010 OR 2011 OR 2012 OR 2013 OR 2014 OR 2015 OR 2016 OR 2017 OR 2018 OR 2019 OR 2020 OR 2021 OR 2022 OR 2023)<br><br>**CABI Digital Library**<br>(Zoono* OR epidemic* OR "disease transmission" OR "disease spread" OR "disease cluster" OR "disease clusters" OR "cluster of cases" OR "animal human" OR "animals human" OR "animal humans" OR "animals humans" OR "infectious disease outbreak" OR "infectious disease outbreaks" OR "infectious diseases outbreak" OR "infectious diseases outbreaks" OR "emerging disease" OR "emerging diseases" OR "reemerging disease" OR "reemerging diseases" OR spillover* OR animal* OR livestock* OR "Crimean-congo hemorrhagic fever" OR "Crimean-congo hemorrhagic fevers" OR "Crimean-Congo haemorrhagic fever" OR "Crimean-Congo haemorrhagic fevers" OR CCHF OR "congo virus" OR "congo viruses" OR "Crimean hemorrhagic fever" OR "Crimean hemorrhagic fevers" OR Marburg OR MVD OR "Middle East |

| | |
|---|---|
| | Respiratory syndrome coronavirus" OR "MERS-CoV" OR "MERS Virus" OR "MERS Viruses" OR "Middle East respiratory syndrome" OR Merbecovirus*) OR (viral AND ("biological threat" OR "biological threats" OR "bio-threat" OR "bio-threats"))<br>AND<br>("East Africa" OR "Eastern Africa" OR "East African" OR "Eastern African" OR Burundi* OR Comoros* OR Djibouti* OR Eritrea* OR Ethiopia* OR Kenya* OR Madagascar* OR Mauritius* OR Rwanda* OR Seychelles* OR Somalia* OR Sudan* OR Tanzania* OR Uganda* OR "Addis Ababa" OR Antananarivo OR Asmara OR Bujumbura OR "Dar es Salaam" OR "Dire Dawa" OR "Djibouti City" OR Dodoma OR Gitega OR Hargeisa OR Juba OR Kampala OR Keren OR Khartoum OR Kigali OR Kisumu OR Lindi OR Mahajanga OR Mbarara OR Mogadishu OR Mombasa OR Muhanga OR Moroni OR Muyinga OR Mwanza OR Nairobi OR Ngozi OR Omdurman OR "Port Louis" OR Victoria OR "Great Lakes Region" OR "Great Lakes Regions" OR "Horn of Africa" OR "Nile Valle")<br>AND<br>(ecosystem* OR ecological* OR climate* OR "natural resources" OR "disaster plan" OR "disaster plans" OR "disaster planning" OR "natural disaster" OR "natural disasters" OR "environmental health" OR "environmental protection") OR ((environment*) AND (drivers OR factors))<br>AND<br>("Policy change" OR "Policy changes" OR "Policy problem" OR "Policy problems" OR "Policy forecast" OR "Policy forecasts" OR "Policy forecasting" OR "Policy solution" OR "Policy solutions" OR "Policy study" OR "Policy studies" OR "Policy environment" OR "Policy environments" OR "Policy development" OR "Policy developments" OR "Policy option" OR "Policy options" OR "Policy alternative" OR "Policy alternatives" OR "Policy recommendation" OR "Policy recommendations" OR "policy implementation" OR "Policy maker" OR "Policy makers" OR "Decision maker" OR "Decision makers" OR "National government" OR "National governments" OR "Policy adoption" OR "Policy evaluation" OR "Policy evaluations" OR "Policy outcome" OR "Policy outcomes" OR "Policy performance" OR "Health policy and planning" OR "policy analysis" OR "Policy formulation" OR "Policy formulations" OR "Policy brief" OR "Policy briefs" OR "Policy reform" OR "Policy reforms" OR "Public policy" OR "public policies" OR "Stakeholder engagement" OR "Stakeholder engagements" OR "Health policy" OR "Health policies" OR "Evidence-based policy" OR "Evidence-based policies" OR "Evidence-informed policy" OR "Evidence-informed policies" OR "risk informed decision making" OR "government authority" OR "government authorities" OR "health planning" OR "Health care reform" OR "Health care reforms" OR Regulation* OR "Land management" OR "Land policy" OR "land policies" OR "Policy engagement" OR "Policy environment" OR "Policy environments") OR ("cross sectoral" AND "decision making") |

| | |
|---|---|
| | **CAB Direct**<br>(Zoono* OR epidemic* OR "disease transmission" OR "disease spread" OR "disease cluster*" OR "cluster of cases" OR "animal* human*" OR "infectious disease* outbreak*" OR "emerging disease*" OR "reemerging disease*" OR spillover* OR animal* OR livestock* OR (viral AND ("biological threat*" OR "bio-threat*")) OR ("Crimean-congo hemorrhagic fever*" OR "Crimean-Congo haemorrhagic fever*" OR CCHF OR "congo virus*" OR "Crimean hemorrhagic fever*" OR Marburg OR MVD OR "Middle East Respiratory syndrome coronavirus" OR "MERS-CoV" OR "MERS Virus*" OR "Middle East respiratory syndrome" OR Merbecovirus*)<br>AND<br>("East* Africa*" OR "Africa* East*" OR Burundi* OR Comoros* OR Djibouti* OR Eritrea* OR Ethiopia* OR Kenya* OR Madagascar* OR Mauritius* OR Rwanda* OR Seychelles* OR Somalia* OR Sudan* OR Tanzania* OR Uganda* OR "Addis Ababa" OR Antananarivo OR Asmara OR Bujumbura OR "Dar es Salaam" OR "Dire Dawa" OR "Djibouti City" OR Dodoma OR Gitega OR Hargeisa OR Juba OR Kampala OR Keren OR Khartoum OR Kigali OR Kisumu OR Lindi OR Mahajanga OR Mbarara OR Mogadishu OR Mombasa OR Muhanga OR Moroni OR Muyinga OR Mwanza OR Nairobi OR Ngozi OR Omdurman OR "Port Louis" OR Victoria OR "Great Lakes Region*" OR "Horn of Africa*" OR "Nile Valle*")<br>AND<br>(ecosystem* OR ecological* OR climate* OR "natural resources" OR "disaster plan*" OR "natural disaster*" OR "environmental health" OR "environmental protection") OR ((environment*) AND (drivers OR factors))<br>AND<br>("Policy change*" OR "Policy problem*" OR "Policy forecast*" OR "Policy solution*" OR "Policy stud*" OR "Policy environment*" OR "Policy development*" OR "Policy option*" OR "Policy alternative*" OR "Policy recommendation*" OR "policy implement*" OR "Policy maker*" OR "Decision maker*" OR "National government*" OR "Policy adoption" OR "Policy evaluation*" OR "Policy outcome*" OR "Policy performance" OR "Health policy and planning" OR "policy analysis" OR "Policy formulation*" OR "Policy brief*" OR "Policy reform*" OR "Public policy" OR "public policies" OR "Stakeholder engagement*" OR "Health polic*" OR "Evidence-based policy" OR "Evidence-based policies*" OR "Evidence-informed policy" OR "Evidence-informed policies" OR ("cross sectoral" AND "decision making") OR "risk informed decision making" OR "government authorit*" OR "health planning" OR "Health care reform*" OR Regulation* OR "Land management" OR "Land policy" OR "land policies" OR "Policy engagement" OR "Policy environment*") |

*Supplemental Table 3 (ST3): List of articles utilized for the critical appraisal of evidence*

| Title | Authors | Published Year | Covidence # | Study |
|---|---|---|---|---|
| 1. **Contribution Of The MESs-CoV Research to One Health Operationalization in Ethiopia and Kenya** | Kiambi, S.; Walelign, E.; Nyariki, T.; Van 't Klooster, G.; Sitawa, R.; Kimutai, J.; Kivaria, F.; Kuria, W.; Njogu, G.; Jobre, Y.; Tewolde, N.; Gari, G.; Bebay, C.; Von Dobschuetz, S.; Gardner, E. | 2022 | #502 | Kiambi 2022 |
| 2. **Short report on implications of COVID-19 and emerging zoonotic infectious diseases for pastoralists and Africa.** | Egeru A; Dejene SW; Siya A | 2020 | #122 | Egeru 2020 |
| 3. **Dromedary Camels: Growing zoonotic disease risk at the human-livestock-wildlife interface** | Zhu, S.; Zimmerman, D.; Deem, S. | 2018 | #633 | Zhu 2018 |
| 4. **Towards a Sustainable One Health Approach to Crimean-Congo Hemorrhagic Fever Prevention: Focus Areas and Gaps in Knowledge.** | Sorvillo TE; Rodriguez SE; Hudson P; Carey M; Rodriguez LL; Spiropoulou CF; Bird BH; Spengler JR; Bente DA | 2020 | #119 | Sorvillo 2020 |
| 5. **The importance of a One Health approach for prioritizing zoonotic diseases to focus on capacity-building efforts in Uganda** | Nantima, N; Ilukor, J; Kaboyo, W; Ademun, ARO; Muwanguzi, D; Sekamatte, M; Sentumbwe, J; Monje, F; Bwire, G | 2019 | #1417 | Nantima 2019 |
| 6. **Zoonotic Pathogens of Dromedary Camels in Kenya: A Systematized Review.** | Hughes EC; Anderson NE | 2020 | #118 | Hughes 2020 |

| | | | | |
|---|---|---|---|---|
| **7. Uganda mountain community health system—perspectives and capacities towards emerging infectious disease surveillance** | Siya, A.; Mafigiri, R.; Migisha, R.; Kading, R.C. | 2021 | #514 | Siya 2021 |
| **8. A descriptive study of zoonotic disease risk at the human-wildlife interface in a biodiversity hot spot in Southwestern Uganda.** | Namusisi S; Mahero M; Travis D; Pelican K; Robertson C; Mugisha L | 2021 | #101 | Namusisi 2021 |
| **9. Lowland grazing and Marburg virus disease (MVD) outbreak in Kween district, Eastern Uganda.** | Siya A; Bazeyo W; Tuhebwe D; Tumwine G; Ezama A; Manirakiza L; Kugonza DR; Rwego IB | 2019 | #173 | Siya 2019 |
| **10. Local, national, and regional viral haemorrhagic fever pandemic potential in Africa: a multistage analysis.** | Pigott DM; Deshpande A; Letourneau I; Morozoff C; Reiner RC Jr; Kraemer MUG; Brent SE; Bogoch II; Khan K; Biehl MH; Burstein R; Earl L; Fullman N; Messina JP; Mylne AQN; Moyes CL; Shearer FM; Bhatt S; Brady OJ; Gething PW; Weiss DJ; Tatem AJ; Caley L; De Groeve T; Vernaccini L; Golding N; Horby P; Kuhn JH; Laney SJ; Ng E; Piot P; Sankoh O; Murray CJL; Hay SI | 2017 | #212 | Pigott 2017 |
| **11. Uganda Tourism Sector COVID-19 Response, Recovery and Sustainability Strategies: Lessons from Previous Virus Disease Outbreaks** | Francis, M.; Jim, A.; Joseph, O. | 2021 | #907 | Francis 2021 |
| **12. Effects of environmental change on zoonotic disease risk: an ecological primer.** | Estrada-Peña A; Ostfeld RS; Peterson AT; Poulin R; de la Fuente J | 2014 | #2788 | Estrada-Peña 2014 |
| **13. A Coupled Human and Natural Systems Framework to Characterize Emerging Infectious** | Manes C; Carthy RR; Hull V | 2023 | #1529 | Manes 2023 |

| | | | | |
|---|---|---|---|---|
| **Diseases-The Case of Fibropapillomatosis in Marine Turtles.** | | | | |
| **14. Preparedness for emerging infectious diseases: pathways from anticipation to action.** | Brookes VJ; Hernández-Jover M; Black PF; Ward MP | 2015 | #2720 | Brookes 2015 |
| **15. Global and regional governance of One Health and implications for global health security.** | Elnaiem A; Mohamed-Ahmed O; Zumla A; Mecaskey J; Charron N; Abakar MF; Raji T; Bahalim A; Manikam L; Risk O; Okereke E; Squires N; Nkengasong J; Rüegg SR; Abdel Hamid MM; Osman AY; Kapata N; Alders R; Heymann DL; Kock R; Dar O | 2023 | #1596 | Elnaiem 2023 |
| **16. A One Health-based Conceptual Framework for comprehensive and coordinated prevention and preparedness to health threats** | Dente, M.G.; Riccardo, F.; Milano, A.; Robbiati, C.; Agrimi, U.; Morabito, S.; Carere, M.; Marcheggiani, S.; Mantovani, A.; Mancini, L.; Villa, L.; Monaco, M.; Scavia, G.; Cubadda, F.; Declich, S. | 2022 | #3994 | Dente 2022 |
| **17. The role of ecosystems in mitigation and management of COVID-19 and other zoonoses.** | Everard M; Johnston P; Santillo D; Staddon C | 2020 | #2117 | Everard 2020 |
| **18. Advances and Limitations of Disease Biogeography Using Ecological Niche Modeling.** | Escobar LE; Craft ME | 2016 | #2561 | Escobar 2016 |
| **19. Grappling with (re)-emerging infectious zoonoses: Risk assessment, mitigation framework, and future directions** | Gwenzi, W.; Skirmuntt, E.C.; Musvuugwa, T.; Teta, C.; Halabowski, D.; Rzymski, P. | 2022 | #6637 | Gwenzi 2022 |
| **20. Use of meteorological data in biosecurity.** | Hemming D; Macneill K | 2020 | #2069 | Hemming 2020 |

| | | | | |
|---|---|---|---|---|
| **21. Data and tools to integrate climate and environmental information into public health.** | Ceccato P; Ramirez B; Manyangadze T; Gwakisa P; Thomson MC | 2018 | #10911 | Ceccato 2018 |
| **22. Eco-social processes influencing infectious disease emergence and spread.** | Jones BA; Betson M; Pfeiffer DU | 2017 | #2560 | Jones 2017 |
| **23. What needs to be done to control the spread of Middle East respiratory syndrome coronavirus?** | Edelstein, M.; Heymann, D.L. | 2015 | #5517 | Edelstein 2015 |
| **24. Filovirus serosurvey following an outbreak of marburg hemorrhagic fever - Ibanda and Kamwenge Districts, Uganda, 2007** | Farnon, E.C.; Adjemian, J.A.; Kansiime, E.; Rahman, J.E.; Bwire, G.S.; Kahirita, S.; Kagirita, A.; Wamala, J.F.; Rollin, P.E. | 2009 | #860 | Farnon 2009 |
| **25. Process Review for Development of Quantitative Risk Analyses for Transboundary Animal Disease to Pathogen-Free Territories.** | Miller J; Burton K; Fund J; Self A | 2017 | #2454 | Miller 2017 |
| **26. Vulnerability assessment tools for infectious threats and antimicrobial resistance: a scoping review protocol.** | Jeleff M; Lehner L; Giles-Vernick T; Dückers MLA; Napier AD; Jirovsky E; Kutalek R | 2019 | #2196 | Jeleff 2019 |
| **27. GIS and Remote Sensing for Malaria Risk Mapping, Ethiopia** | Ahmed, A | 2014 | #14115 | Ahmed 2014 |
| **28. Reemergence of Marburgvirus disease: Update on current control and prevention measures and review of the literature.** | Elsheikh R; Makram AM; Selim H; Nguyen D; Le TTT; Tran VP; Elaziz Khader SA; Huy NT | 2023 | #3 | Elsheikh 2023 |
| **29. Decision-support tools to build climate resilience against** | Rocklöv, J.; Semenza, J. C.; Dasgupta, S.; Robinson, E. J. Z.; Abd El Wahed, A.; Alcayna, T.; | 2023 | #10734 | Rocklöv 2023 |

| | | | | |
|---|---|---|---|---|
| **emerging infectious diseases in Europe and beyond** | Arnés-Sanz, C.; Bailey, M.; Bärnighausen, T.; Bartumeus, F.; Borrell, C.; Bouwer, L. M.; Bretonnière, P. A.; Bunker, A.; Chavardes, C.; van Daalen, K. R.; Encarnação, J.; González-Reviriego, N.; Guo, J.; Johnson, K.; Koopmans, M. P. G.; Máñez Costa, M.; Michaelakis, A.; Montalvo, T.; Omazic, A.; Palmer, J. R. B.; Preet, R.; Romanello, M.; Shafiul Alam, M.; Sikkema, R. S.; Terrado, M.; Treskova, M.; Urquiza, D.; Lowe, R. | | | |
| **30. Advancing One human-animal-environment Health for global health security: what does the evidence say?** | Zinsstag J; Kaiser-Grolimund A; Heitz-Tokpa K; Sreedharan R; Lubroth J; Caya F; Stone M; Brown H; Bonfoh B; Dobell E; Morgan D; Homaira N; Kock R; Hattendorf J; Crump L; Mauti S; Del Rio Vilas V; Saikat S; Zumla A; Heymann D; Dar O; de la Rocque S | 2023 | #1598 | Zinsstag 2023 |
| **31. Tracking the distribution and impacts of diseases with biological records and distribution modelling** | Purse, B.V.; Golding, N. | 2015 | #7150 | Purse 2015 |
| **32. Mapping Potential Amplification and Transmission Hotspots for MERS-CoV, Kenya.** | Gikonyo S; Kimani T; Matere J; Kimutai J; Kiambi SG; Bitek AO; Juma Ngeiywa KJZ; Makonnen YJ; Tripodi A; Morzaria S; Lubroth J; Rugalema G; Fasina FO | 2018 | #198 | Gikonyo 2018 |

*Supplemental Table 4 (ST4): Environmental drivers and pathogen transmission routes*

| Environmental Driver | Pathogen | Transmission Route | Linkage to climate change | Examples of cited evidence (epidemiological, ecological, and/or serological) | Literature Reference Source examples |
|---|---|---|---|---|---|
| **1. Climate Change**[3,4,24,33–55] | Crimean-Congo haemorrhagic fever virus (CCHFV), Middle East respiratory syndrome coronavirus (MERS-CoV), Marburg virus (MARV) | - Increase in feeding frequency of vectors like CCHFV-infected ticks due to rising temperatures<br>- Population migration and herd mixing near wildlife watering points increases potential of spillover events from MERS-CoV reservoir hosts (dromedary camels) to humans and/or among livestock.<br>- Changes in wind pattern facilitates condition for pathogen dispersal across geographic boundaries<br>- Ecological imbalance caused by climate change results in mixture of different specie/viral strains and cross-species disease transmission.[41] | n/a | - Dry season reported as a major risk factor for exposure to CCHFV in Kenya (OR: 7.197; 95% CI: 0.813 – 63.65).[37]<br>- Highest CCHFV seropositivity reported in camels in central Sudan residing in a locality characterized by dry climate and temperature variations.[4]<br>- Climate change, and climate-induced drought and desertification reported as risk factors for tick distribution in Sudan.[38]<br>- Climate change and land use change (due to climate-related expansion of arid regions and increase in pastoralism) were found to be spatially correlated with MERS-CoV cases in Kenya from 2016-2018.[43]<br>- Findings from ecological niche modeling (ENM) studies report influence of climate change on geological distribution of MARV | - Lawrence et al. 2024<br>- Elsheikh et al. 2023<br>- Blanco-Penedo et al. 2021<br>- Shuaib et al. 2020<br>- Suliman et al. 2017<br>- Peterson et al. 2016<br>- Kimaro et al. 2013<br>- Peterson et al. 2006 |

| | | | | | |
|---|---|---|---|---|---|
| | | | | (correlated with seasonal temperature variations).[36,39,40] | |
| **2. Migration Patterns (Human, animals, wildlife).**[4,6,20,25,28,37,41,48,49,51,56–78] | MERS-CoV, CCHFV, MARV | - Migratory birds in search of new favorable habitat transport Crimean-Congo haemorrhagic fever (CCHF)-infected ticks to non-endemic areas<br>- MARV-infected Fruit bats migration from natural reservoirs to new roosting and foraging sites increases spillover risk upon human and livestock contact with infected bats drooping<br>- Climate change induced migration of MERS-CoV infected dromedary camel and other livestock to communal watering points/grazing land facilitates cross-species pathogen transmission<br>- Transboundary movement of people and livestock along trade routes | Impacted by climate change with an increase in human displacement and non-human species movement | - Findings from knowledge, attitudes, and practices (KAP) survey in Garissa and Isiolo counties, Kenya on migration of camels to communal watering points, grazing areas and marketing points as high risk driver of the spread of MERS-CoV.[57]<br>- Findings from a seroprevalence study in cattle (31% seropositivity): Population and livestock migration patterns from arid to semi-arid regions of Kenya to riverine areas facilitate cross-species transfer of pathogens from one region to another.[37]<br>- Livestock movement associated with CCHFV seroprevalence in Madagascar.[6]<br>- Mass migration of livestock across Kenya in search of grazing land increases potential for dispersal of tick-borne CCHFV.[72]<br>- Highest CCHF prevalence reported in Tana River county associated with formation of flood plains | - Omoga et al. 2023<br>- Lule et al. 2022<br>- Othieno et al. 2022<br>- Chiuya et al. 2021<br>- Blanco-Penedo et al. 2021<br>- Roess et al. 2015<br>- Tigoi et al. 2015<br>- Corman et al. 2014<br>- Andriamandiby et al. 2011 |

| | | | | | |
|---|---|---|---|---|---|
| | | | | during floods and increased temperature favorable to disease transmission; viral spread is further exacerbated by convergence of different species for water and pasture during dry season.[60]<br>- Livestock movement associated with high CCHFV exposure risk in Uganda and Kenya;[56,58] and MERS-CoV exposure risk in Ethiopia[59] and across East Africa.[64,67]<br>- Increase in MERS seropositivity from Western Kenya to Northeastern and eastern regions associated with increase in cross-border nomadic dromedary camel population.[61]<br>- Recent study reported the. First evidence of CCHFV seropositivity among rodents (in addition to livestock and humans) in Kenya [6.9%: (6/93)].[20] | |
| **3. Habitat Encroachment** [11–15,36,42,47,72,78–119] | MARV, MERS-CoV | - Pathogen spillover from reservoirs hosts (Fruit bats) roosting along the periphery of caves and mines to humans in contact with | Contributes to | - Significant association between previous mining activity and prior MARV infection (OR: 25.1; 95% CI 5.4 – 118).[91]<br>- Miners and family members have 2.2 times the risk of | - Rugarabamu et al. 2022<br>- Rugarabamu et al. 2021<br>- Nyakarahuka et al. 2020 |

| | | | | | |
|---|---|---|---|---|---|
| | | fruit bats and/or bat secretions | | filovirus infection (RR: 2.2; 95 CI 0.98 – 4.7).[90]<br>- Visits to mines and caves inhabited by fruit bats (RR:2.5; 95% CI: 1.12 – 5.66).[90]<br>- Miners in Western Uganda have 5.4 times the risk of filovirus seropositivity compared to non-miners in Central Uganda (RR: 5.4; 95% CI: 1.5 – 19.7).[89]<br>- Association between mining and filovirus seropositivity (AOR: 3.4; 95% CI: 1.3 – 8.5); and entering mines and filovirus seropositivity (AOR: 3.1; 95% ci: 1.2 – 8.2).[89]<br>- Association between Marburg virus disease (MVD) seropositivity and contact with cave roosting bats (OR: 1.2; 95% CI: 1.1 – 1.5).[87,88]<br>- Linkage of frequent salt mining and manure collection activities by livestock keepers, during visits to caves inhabited by Rousettus spp. Bats, as a primary risk factor for spillover of MARV to humans.[12,92] | - Nyakarahuka et al. 2017<br>- Farnon et al. 2009)<br>- Adjemian et al. 2011<br>- Sang & Dunster, 2001 |

| | | | | | |
|---|---|---|---|---|---|
| 4. **Observed Ecological changes due to agricultural intensification** [30,34,43,52,56,118,120–123] | CCHFV, MERS-CoV, MARV | Transmission from *Hyalommma marginatum* and *Rhipicephalus appendiculatus* tick species | Contributes to [associated with increases in greenhouse gas (GHG) emissions) for e.g., poultry production requires use of fossil fuels, and manure use and storage results in emission of GHGs. (Drozdz et al., 2020) and impacted by (increase in extreme weather events results in intensified agricultural practices to improve crop yields). | - Based on eco-epidemiological study, significant predictor of tick transmission in Uganda reported to be agricultural land use, vegetation seasonality (NDVI mean and seasonality), and precipitation and temperature factors; high environmental suitability for CCHFV-infected *R. appendiculatus* tick.[56]<br>- CCHFV seroprevalence of 10.3% (7-8-13.3) in humans; 69·7% (65·1–73·4) in cattle[56]<br>- Significant association between CCHFV seroprevalence and proximity to agricultural land; compared to anthropogenic variables, inclusion of environmental variables like enhanced vegetation index, land surface temperature and percent sand-soil composition were better predictors of CCHFV seroprevalence among livestock (31.4%).[121]<br>- 90% MERS-CoV seroprevalence among | - Telford et al. 2023<br>- Lule et al. 2022<br>- Magenas et al. (2022)<br>- Munyua et al. 2017 |

| | | | | | |
|---|---|---|---|---|---|
| | | | | camel population in Kenya (100% in Marsabit county); Ecological changes due to intensive agricultural practices increases risk of MERS-CoV transmission to humans.[30] | |
| **5. Deforestation and Reforestation**[36,72,124] | MARV | Impacts of deforestation due to mining and logging activities results in the redistribution of bats habitat and facilitate MVD emergence and spread to non-endemic areas | Contributes to climate change | - ENM and species distribution modeling (SDM studies) reported a significant correlation between biological reservoirs including filoviruses and deforestation (AUC: 0.978).[124] | - Jagadesh et al. 2020 |
| **6. Animal Husbandry practices**[1,5,18,19,21,29,47,49,61,70,71,74,125–133] | CCHFV, MERS-CoV | Herd mixing of livestock with disease infected wildlife; increased interaction of immunologically naïve and infected animals during transportation of livestock from different herds and in holding pens[125] | Contributes to | - Higher MERS-CoV seroprevalence in camels raised on pastoral production system (69%; 95% CI: 66.5 - 71.6) compared to ranching production system (10.1%; 95% CI: 4.9 – 15.3).[74,126]<br>- Cattle raised on open grazing system reported to be 27 times more at risk of CCHFV infection compared to cattle raised in closed grazing system (OR: 27.22; 95% CI: 7.46- 99.24).[5]<br>- Association between poor animal husbandry of famers in contact with donkeys and | - Ogoti et al. 2024<br>- Obanda et al. 2021<br>- Sitawa et al. 2020<br>- Gardner et al. 2019<br>- Ibrahim et al. 2015<br>- Lwande et al. 2012 |

| | | | | | |
|---|---|---|---|---|---|
| | | | | CCHFV exposure (OR: 1.92; 95% CI: 1.05 - 3.72).[21]<br>- 95% CCHFV seroprevalence rate reported in African buffalo raised in closed wildlife grazing systems in Kenya compared to seropositivity rates of 29% and 46% reported for closed integrated and open integrated grazing systems respectively.[19] | |
| 7. **Livestock Overgrazing**[2,134,135] | CCHFV, MARV | Expansion of livestock grazing lands to overlap into areas typically inhabited by wildlife provides more opportunity for cross-species pathogen transmission | Contributes to | - People living close to livestock grazing fields reported as 18 times more likely to be CCHF case patients than controls (OR: 18, 95% CI: 3.2 - ∞)<br>- Cross sectional study and focus group discussion focused on community perspective reported lowland grazing, settling in caves during we season, and salt mining and utilization of bat droppings as livestock fertilizer as major risk factors for MVD emergence in Uganda[135] | - Mirembe et al. 2021<br>- Siya et al. 2019 |
| 8. **Biodiversity loss**[42,49,104,136] | CCHFV | - Expansion of livestock grazing land due to climate change results in biodiversity loss | Impacted by | - Reported declines in vulture population linked to rise in diseases spread via abattoirs in Eastern Africa including tick-borne diseases.[136] | - Rodarte et al. 2023<br>- Amman et al. 2023 |

| | | | | | |
|---|---|---|---|---|---|
| | | including loss of species serving basic ecosystem function (for e.g. disease control).<br>- land use changed linked to increase in zoonotic reservoir host species and a decline in non-reservoir species/wildlife population<br>- Declines in vulture population around abattoirs increase opportunities for pathogen transmission from infected ticks feeding on livestock in abattoirs | | - Findings from micro-global positioning study support earlier evidence about decrease in non-reservoir species due to land use change | |
| **9. Irrigation Practices**[72] | CCHFV, MERS-CoV | Contained water sources (independent of seasonal rain fall patterns) prolongs pathogen vector breeding periods and provides more vector breeding habitat that result in increased risk of disease transmission | Impacted by, and contributes | Human settlements around irrigation sites linked to increased risk of CCHFV transmission in Kenya.[72] | Sang & Dunster 2001 |

| | | | | | |
|---|---|---|---|---|---|
| **10. Human-mediated transport of pathogens across geographic boundaries ("pathogen pollution")**[93,125] | MARV, MERS-CoV | Transport of MERS-CoV infected nomadic camels to abattoir hubs increases MERS-CoV exposure risk to abattoir workers | Impacted by, and contributes to | Seasonal factors (such as dry seasom) were associated with increased MERS-CoV RNA positivity in camels [36/2711 (1.3%] and biphasic incidence of MER-CoV infections from nomadic camels to abattoir workers in Northern Kenya [October 2022 (7/60, 11.7%); and February 2023 (7/58, 12.1%)].[125] | Ogoti et al. 2024 |

*Supplemental Note 1 (SN 1): Summary of Risk Frameworks and Areas of Application*

i) Risk Mapping-Based Assessment Tools: Reported application of conventional risk maps include for use as a forecasting system and decision support tool (DST) during the 2006-2007 Rift Valley fever (RVF) outbreaks linked to El-Nino episodes in Kenya;[137] identifying and mapping MERS-CoV transmission hotspots and risk nodes along the camel value chain in Kenya;[63] and anthrax risk mapping to inform risk communication and decision making.[138] Specialized risk mapping approaches like ecological niche modeling (ENM) are reportedly used for filling surveillance data gaps and other gaps in conventional risk mapping approaches. Some studies reported on the application of ENM combined with remote sensing technologies to enhance environmental monitoring data including for flood and other environmental hazard detection in the East Africa region.[139,140] Related to ENM application for environmental monitoring, a study carried out in Uganda found that socio-environmental variables like land cover, average annual change in nighttime light index, EVI, Normalized Difference Vegetation Index (NDVI) and percent sand-soil content have the best predictive accuracy for risk of CCHFV transmission among livestock;[121] and one study described its application for determining environmental and socio-ecological suitability for CCHF infections in Uganda.[56] Another study on the environmental suitability for risk of MERS-CoV transmission found the most significant predictor variables to be bare land coverage, forest coverage, and population density;[141] and a review article highlighted the geological distribution of MARV and its association with climate change (in relation to pathogen distribution across precipitation areas on the African continent) with a reported correlation between MVD and seasonal temperature variations characterized by low to moderate temperature and precipitation.[36,107] ENM also associates temperature seasonality, rainfall, and vegetation indices as key determinants of the spatial distribution of MARV.[39,107,115,142,143]

ii) *Multivariable risk assessment and management frameworks*: Included in the review findings are risk assessment and risk management frameworks that incorporate a combination of different categories of variables for multihazard risk assessment. These frameworks include the Index For Risk Management Epidemic Risk Index (INFORM ERI), an open-source risk assessment tool adopted from the Index for Risk Management Global Risk Index (INFORM GRI) and used to address gaps in research on risks of climate-related hazards and exposures including epidemic risks of four viral hemorrhagic fevers of epidemic potential;[144,145] Spatial Multicriteria Decision Analysis tool utilized for assessing emergence of West Nile Virus in China;[146] WHO Health Emergency and Disaster Risk Management (EDRM) framework that utilizes an all-hazards risk management approach to mitigating the health risks and consequences of emergencies including climate-induced infectious disease outbreaks;[147] UNDRR Technical guidance framework on application of climate information for comprehensive risk management;[148] and integrated risk and vulnerability assessment framework developed for assessing the risk of climate change associated malaria transmission at the community level in East African highlands.[149]

iii) *Qualitative and semi-quantitative risk tools based on expert opinions*: Some studies reported the use of expert-informed qualitative multisectoral risk assessment tools such as the Tripartite Joint Risk Assessment Operational Tool ( JRA OT) used for addressing zoonotic biological threats at the animal-human-environment interface across different countries including for the risk assessment of rabies and influenza in Jordan,[150] and as part of the OH zoonotic disease prioritization exercise in Ukraine.[151] Reported limitations of the tool include real-time cross-sectoral data gaps due to gaps in surveillance systems across the country thus contributing to a higher level of uncertainty associated with the Joint Risk Assessment (JRA) process and the need to strengthen the environment-interface by better integrating the environment sector into the JRA process.[150]

Other risk assessment tools informed by expert opinions include hybrid risk ranking tools such as the Spillover Viral Risk Ranking tool used to assess the spillover and pandemic potential of zoonotic viruses,[152] to track potential reservoirs of emerging pathogens[153] and to assist countries with priority zoonotic diseases risk ranking efforts to inform resource allocation;[154] infectious disease seeker software tool used for predicting, assessing, and comparing different outbreaks;[139,155] a risk classification methodological tool used for classifying influenza viruses;[156] semi-quantitative risk assessment tool utilized for the risk assessment of spillover routes and disease amplification;[157] and the Africa CDC Risk Classification and Prioritization of Epidemic-Prone Diseases tool to identify priority diseases of epidemic potential to inform resource allocation and implementation of risk mitigation measures.[158]

iv) *Quantitative-based environmental risk assessment tools:* Application of environmental risk assessment tools utilized for detecting zoonotic biological threats include environmental vulnerability and preparedness risk assessment checklist;[159] Quantitative Microbial Risk Assessment (QMRA) to investigate potential health risks of severe acute respiratory syndrome coronavirus 2 (SARS-CoV-2) for vulnerable groups such as wastewater treatment plant workers and for scenario planning.[160] Another study reported on the application of QMRA in combination with remote sensing technology to monitor cross-species population movement and potential for disease transmission,[161] and for the assessment of pathogens in aquatic environments.[162] Using a similar environmental approach, Bayesian Belief Networks were used to make disease risk estimation, characterization, and prediction for climate adaptation.[163]

*Table 5: Summary of risk frameworks/methodological approaches and areas of application*

| Integrated Risk Framework/ Methodological Approach | Example Description | Unique Features and Benefits | Example areas of application in the region | Extent of Geographic Coverage | Highlighted Gap areas |
|---|---|---|---|---|---|
| **Ecological Niche Modeling (ENM)/ Species Distribution Modeling (SDM)**[36,39,107,115,118,124,139,141,143,145,153,164–176] | Based on environmental interpolations/estimations of potential distributions; useful for identifying at-risk areas by utilizing data on environmental correlation found in areas with strong disease surveillance and reporting coverage to make risk predictions in areas with limited surveillance and reporting and thus little knowledge about disease transmission potential.[166] | - Adopts different types of Algorithms including Maximum Entropy (MaxEnt)[107,141,175], Boosted Regression Trees[56,115], Random Forest, Genetic Algorithm for Rule-set Prediction (GARP)<br>- Uses both presence only occurrence data and absence data to provide an estimation of | - For detection of potential risk of MARV spillover in countries across SSA.[115]<br>- Potential for human rabies transmission in non-endemic area.[167]<br>- Potential outbreaks of filovirus Uganda.[90]<br>- Potential tick host of Crimean-Congo haemorrhagic fever virus (CCHFV) and widespread exposure to the virus in Uganda.[56]<br>- Risk of Middle East respiratory syndrome coronavirus (MERS-CoV) transmission globally.[141] | Global, National, Regional, Local (sub-national) | Need to improve ENM validation systems, and account for gaps in reporting of occurrence data across countries to reduce bias in distribution models |

| | | | | | |
|---|---|---|---|---|---|
| | | both potential distribution (ENM) and actual distribution (SDM) of a species/pathogen<br>- Predicts potential hotpots and suitable environments for future outbreaks helpful for prioritizing resource-limited high-risk areas for surveillance, prevention, preparedness, and control efforts. | | | |
| **Geospatial Analysis and Remote Sensing data systems;**[137,139,140,142,161,163,177–183] **and Spatial Multicriteria decision analysis tool** | Useful for large-scale environmental mapping and surveillance including for the identification of risk factors contributing to multiple clusters of emerging and/or reemerging infectious diseases (EIDs) such as | - EPIDEMIA provides an interdisciplinary approach to malaria surveillance, forecasting and decision support to inform malaria prevention, | Amhara region, Ethiopia<br><br>Northwestern, Eastern, and Southern China<br><br>MERS-CoV seroprevalence in Kenya | National and Sub-national | Need for the development of a comprehensive, open access database for sharing, storing, and accessing remotely-and proximally sensed data sources |

| | | | | | |
|---|---|---|---|---|---|
| **(SpatMCDA)**[138,146,174,184,185] | coronavirus emergence.[182] | control and elimination in Ethiopia.[178]<br>- SpatMCDA is utilized for assessing areas at risk of EIDs such as West Nile virus especially in resource limited settings.[146]<br>- Used to determine the spatial relationship of EIDs to variables such as climate, agriculture and sociodemographic factors.[43] | | | |
| **Traditional Risk Mapping**[137,186–188] | Use of 2-3 dimensional visualizations depicting high vs. no risk areas[138]; Based on disease occurrence density and spatial interpolations | Used as a disease forecasting system to make predictions about areas with risk of disease emergence to inform risk communication efforts | Food and Agriculture Organization (FAO) Rift Valley fever (RVF) Decision support tools for enhancing capacity for early warning and forecasting of RVF in high-risk countries (such as Kenya, Tanzania, Uganda) in the East Africa region.[137,188] | National, Sub-national | Does not account for risk in areas with limited/lack of data on disease occurrence (absence data) due to low surveillance coverage |

| | | | | | |
|---|---|---|---|---|---|
| **Integrated Risk and Vulnerability Assessment**[149] | Conceptual framework utilizing a combination of variables to assess the interplay among biophysical (especially climate change) and socio-economic and cultural factors on malaria transmission in East Africa | Adopts the use of a combination of variables to assess risk of vector-borne disease transmission at the community level | Community level risk of malaria transmission due to climate change in the East Africa region.[149] | Community level | The need for the adoption of a combination of empirical quantitative and qualitative data sources including from communities, and expert opinion, to ensure the development of a more robust risk analysis model for decision making |
| **Joint Risk Assessment Operational Tool (JRA OT)**[150,151,189]; **other multisectoral risk assessment approaches**[190–192] | Useful for collating cross-sectoral expert opinion to characterize disease risk using a mostly qualitative data | Helpful guide for bringing together all relevant sectors to assess shared threats and to inform risk management and risk communication; and resource allocation[193] | Joint risk assessment (JRA) of rabies and Avian influenza in Jordan.[150] | National and Sub-national | - Need to expand the scope of the tool to threats related to environmental hazard and exposures including better utilization of the tool for assessing the impact of environmental hazards on the spillover, emergence, and spread of zoonotic biological threats; better integration of the environmental sector as a key technical partner in the operationalization of the tool; integrating the United Nations Environment Programme (UNEP) and national environmental sector risk assessment processes to the JRA OT |

| | | | | | |
|---|---|---|---|---|---|
| | | | | | operationalization process.<br>- Reported gaps in surveillance systems increase uncertainty level associated with the JRA process<br>- Addressing real-time/near real-time data gaps to better quantify risk level |
| **Index for Risk Management Global Risk Index (INFORM GRI/ Epidemic Risk Index (INFORM ERI)**[144,194] | Used to address gaps on risks of climate-related hazards and exposures in relation to epidemic risks to inform decision-making.[194] | Adapted from the Index for Risk Management Global Risk Index (INFORM GRI) to specifically analyze environmentally driven biological threat exposure risks, population vulnerabilities, and lack of coping capacities to inform the implementation of appropriate risk mitigation and risk communication strategies | Application by Pigot et al. (2017) show how an index case escalated into a widespread epidemic and areas of potential susceptibility at local, regional, and global level.[195] | Global, Regional, National | Need to expand application to diverse range of pathogens with epidemic and pandemic potential |
| **Risk Ranking Tools**[153–158,196] | - SpillOver Viral Risk Ranking Framework: Web-based risk | - Open-source and adaptable platform for | - Can be used in combination with ENM to better | Global, Regional | - Lack of/limited data on wildlife source of animal-human spillover |

| | | | | | |
|---|---|---|---|---|---|
| | assessment tool used to evaluate and assess the zoonotic spillover and pandemic potential of novel viruses.[196]<br>- Infectious Disease seeker software tools: used for predicting, assessing, and comparing different outbreaks.[155]<br>- Risk Ranking and Prioritization of Epidemic-Prone Diseases tool: used for prioritization of diseases of epidemic potential to inform resource allocation and implementation of risk mitigation interventions.[158] | collating data on spillover risk factors and assigns a comparative risk score for viruses of wildlife origins | characterize emerging pathogen reservoir host species | | transmission risk reduces the accuracy of risk ranking tools (resulting in over- or under-estimation of assigned risk score); inconsistencies with reporting on pathogen host species<br>- Data gaps serve as a hindrance to properly characterizing risk factors |

| | | | | | |
|---|---|---|---|---|---|
| **Other environmentally focused risk assessment methodological tools [Quantitative Microbial Risk Assessment (QMRA),**[157,160–162] **Bayesian Networks Modeling]**[124,157,163] | Utilizes a combination of quantitative and qualitative data for environmental monitoring | Combines qualitative expert knowledge with quantitative data including remote sensing data for model development | - Adoption of QMRA to investigate health risks of severe acute respiratory syndrome coronavirus 2 (SARS-CoV-2) to vulnerable groups such as wastewater treatment plant workers.[160]<br>- Use of Bayesian Networks for risk assessment of spillover routes, disease amplification and spread in South Korea.[157] | National | - Data intensive: reported challenges with data gaps in both developed and developing countries<br>- Need to better account for the perspective of stakeholders in model design, implementation, and communication of findings |

*Supplemental Note 1: Implications for public health practice*

Based on the integrative review findings, our proposed integrated risk analysis (IRA) framework primarily incorporates a combination of the ecological, meteorological, biological, and socio-anthropogenic drivers that increase pathogen spillover risk and disease emergence/reemergence into the risk assessment and risk characterization steps. In line with the WHO Health EDRM framework,[197] the IRA framework also addresses the need for proactive health systems resilience building as a risk mitigation strategy to mitigate the impact of environmentally driven biological events of potentially catastrophic magnitude. This new framework also accounts for recommendations for an integrated OH-based conceptual framework informed by expert opinions to facilitate operationalization of OH. Thus, the framework addresses the need to enhance the integration of the environment sector into the OH approach for the implementation of pandemic prevention and preparedness activities including climate-resilient health systems resilience building measures.[190,198–200]

The figure below *(Figure 1*) describes the application of the newly conceptualized integrated risk analysis (IRA) to provide a coordinated cross-sectoral response to a hypothetical cluster of future cases of Crimean-Congo haemorrhagic fever (CCHF) infections in Kenya. The scenario presented is in line with the integrative review findings on past CCHF outbreaks across countries in the East Africa region. The IRA framework incorporates key findings from the review about addressing gaps in existing risk analysis framework and methodological tools to address environmentally driven zoonotic biological threats at the animal-human-environment interface using an integrated approach to risk analysis.

*Figure 1: Application of the newly conceptualized Integrated Risk Analysis framework model*

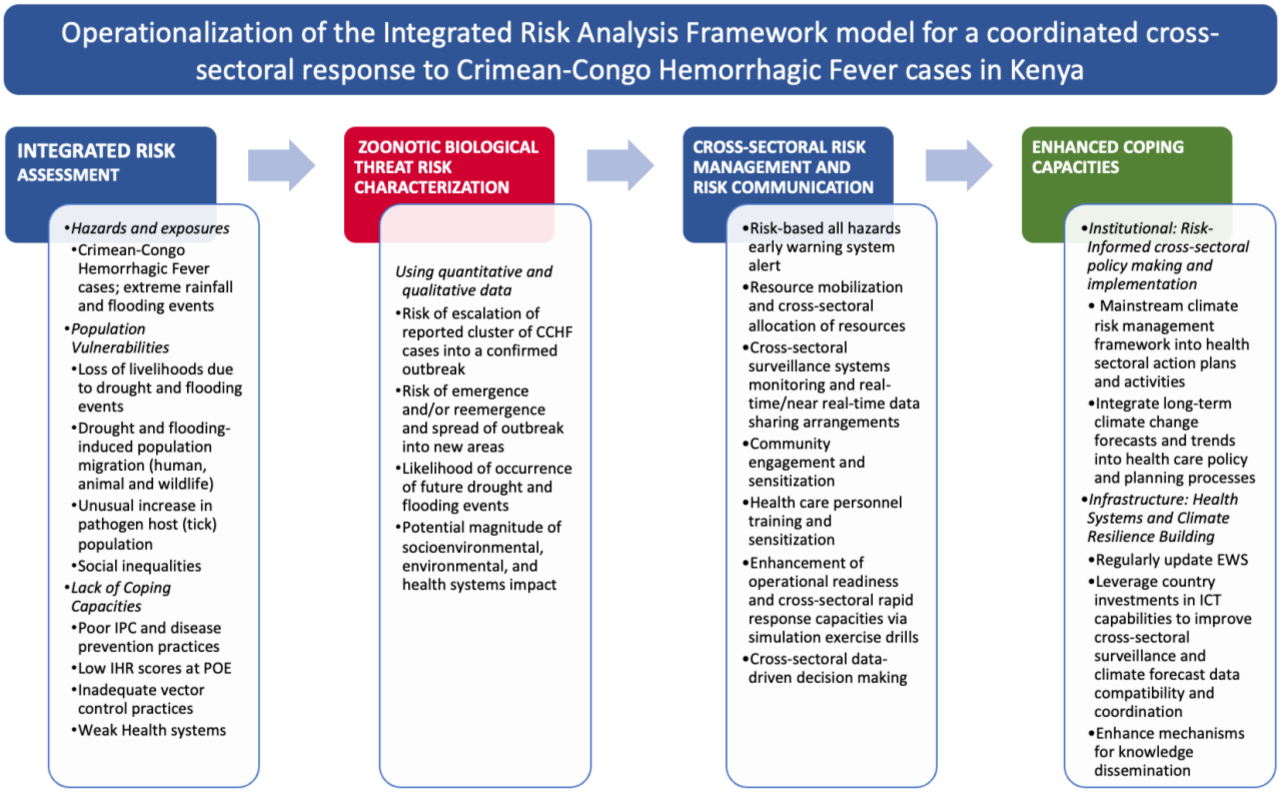

**Box 1 - A Case Study: Operationalization of a newly conceptualized Integrated Risk Analysis (IRA) framework for a coordinated cross-sectoral response to the reemergence of Crimean-Congo Haemorrhagic Fever (CCHF) in two high-risk counties in Kenya**

Converging risk drivers

*Hazards and exposures*

The Ministry of Health (MoH) recently received an alert about suspected cases of CCHF in two counties in Kenya. As an integral part of the cross-sectoral response to the reported cluster of cases, the Ministry of Environment, Climate Change and Forestry closely monitors national climate data and shares information on common climate-related hazards (flooding and drought) events at the national and county levels that are known to have an impact on vector-borne disease transmission patterns. The risk analysis also considers past trends and future projections about climate change and climate-related multihazard occurrence, based on ecological niche modeling (ENM)/species distribution modeling (SDM) data, climate models, and geospatial and remotely sensed data. This information is included in the analysis of epidemiological surveillance data from the MoH and the Ministry of Agriculture and Livestock Development (MALD) on suspected and confirmed cases of CCHF in animals and humans.

*Population vulnerabilities*

In both affected counties and cross-border counties with no known reported case of CCHF, receding lake levels, drying of the rivers and other wetlands puts local communities that rely on fishing and agriculture as a primary source of income in a more vulnerable state. With the approaching drought season, there has been mass population migration from the arid and semi-arid zones of the country towards the highlands to escape the socioeconomic impact of droughts due to agriculture and livestock loss. Similarly, past incidences of flooding events due to extreme rainfalls are causing frequent land degradation, soil erosion and water logging of crops which result in reduced crop yields, increased food insecurity, and an unusual increase in tick population. In addition to community displacement, the effects of these climate-related hazards provide an economic incentive for mass human, animal, and wildlife population migration to other areas in search of new habitat. Hence, available data on changing migration patterns, economic development, and social inequality will be accounted for in the risk assessment.

*Lack of coping capacities*

At the devolved county government levels, poor infection, prevention and control (IPC) and disease prevention practices, and low levels of International Health Regulations (IHR) core capacities scores at points of entry have been identified as key weaknesses in coping capacities. In addition, underlying causes of vulnerabilities include gaps in water and sanitation services, weaknesses in health systems capacity to address climate-related hazards, poorly implemented veterinary regulations, as reported by the MALD, and inadequate vector control practices.

Risk characterization

Based on the initial assessment of available cross-sectoral data, the risk characterization accounts for the level of CCHF exposure, population vulnerabilities, and capacities at national and county level. Exposure levels consider the risk of escalation of reported clusters of cases into a confirmed outbreak, and the emergence/reemergence of the outbreak into new areas. Additionally, the likelihood of occurrence of future climate-related hazards (droughts and floods) and the potential magnitude of impact in terms of socioeconomic, environmental, and health systems impact is also included in the risk characterization.

**Box 1 (continued) - A Case Study: Operationalization of a newly conceptualized IRA framework for a coordinated cross-sectoral response to the reemergence of CCHF in two high-risk counties in Kenya**

Cross-sectoral Risk Management and Risk Communication

Following a heightened situational awareness about the reported clusters of CCHF cases, an early warning system (EWS) is triggered to indicate the need to take concrete actions (specific to the local context) including targeted mobilization and allocation of resources across different line ministries to enable further investigations, close monitoring of surveillance systems and cross-sectoral sharing of data in real time/near-real time to further update the risk assessment as more information becomes available. In addition, community engagement, sensitization and surveillance of unusual events is prioritized to alert health authorities and to ensure inclusion of the most-affected populations in the risk management decision-making process. As part of current risk mitigation and future risk reduction efforts, resources are also mobilized for improving health systems preparedness capacity including for the sensitization and training of health care personnel on improving IPC practices, while ensuring safe treatment of identified cases, and for the monitoring and prevention of climate-related diseases. Plans are also put in place to enhance operational readiness and rapid response capacity via simulation drills in preparation to respond to large-scale outbreaks and other climate-related hazards. This involves the use of ENM/SDM, climate modeling, remotely sensed data, and surveillance data on environmental drivers for data-driven decision making and awareness raising to ensure increased political commitment and sustained investments towards prevention of climate-related hazards and future preparedness planning using a OH approach.

Outlook: Enhanced Coping Capacities

The actions to be taken to enhance capacities address underlying risk drivers of CCHF emergence/reemergence and spread in affected counties, while focusing on future preparedness, mitigation, and long-term resilience building efforts. Proactive actions for strengthening coping capacities include regularly updating EWS based on most recent data on environmental drivers of EIDs to trigger a faster response to future outbreaks; improving the political, legal, and regulatory environment to ensure the adoption of effective OH policies (including climate-smart agriculture policies and integration of long-term climate change forecasts and trends into health care policy and planning processes); and implementation of OH risk-based activities under existing policies such as the National Action Plan for Health Security (NAPHS) and country environment action plan. Such activities include integrating cross-sectoral surveillance data and enabling better use of environmental, climate change, public health, and animal health data at the national and county level, mainstreaming climate risk management into health sectoral plans and activities, in line with Kenya's Integrated Climate Risk Management Framework, leveraging on the country's investments in Information Communication and Technology (ICT), meeting commitments towards improving cross-sectoral knowledge sharing platforms and information management systems to ensure data compatibility and coordination across sectors, and improving mechanisms for community engagement and knowledge dissemination to inform future EIDs risk analysis.
